# Melanocyte loss dominates the vitiligo transcriptome: a rank-based meta-analysis

**DOI:** 10.64898/2026.02.07.26345817

**Authors:** Xijin Ge

## Abstract

Vitiligo is an autoimmune disorder characterized by the destruction of melanocytes. We performed a rank-based meta-analysis of six independent transcriptomic studies (115 samples) spanning microarray, bulk, and single-cell RNA-seq platforms to identify consensus signatures of lesional skin. Robust rank aggregation identified 108 downregulated and 6 upregulated genes. Pathway analysis revealed consistent suppression of melanin synthesis and neural development pathways in vitiligo, whereas immune response activation was heterogeneous across studies. Re-analysis of single-cell data from three studies confirmed melanocyte depletion. The 108 downregulated genes were expressed exclusively in melanocytes. These include neural development genes (PLP1, GPM6B, NRXN3), consistent with melanocytes’ neural crest origin. We also identified candidate melanocyte markers, such as CYB561A3 and QPCT, with high melanocyte specificity and consistent downregulation in vitiligo. These findings reveal a robust melanocyte-loss signature in vitiligo, detectable across different studies. Study-dependent immune activation, possibly influenced by sampling method and disease characteristics, warrants further study.

## Introduction

Vitiligo is an acquired depigmenting disorder affecting 0.5-2% of the global population (Ezzedine et al., 2015), characterized by the selective loss of melanocytes (Frisoli et al., 2020). Active depigmentation involves IFN-γ signaling and the CXCL9/10-CXCR3 chemokine axis, which recruits autoreactive CD8+ T cells to the skin (Harris et al., 2012; Rashighi et al., 2014). In stable lesions, tissue-resident memory CD8+ T cells persist and can be reactivated, preventing repigmentation (Boniface et al., 2018; Richmond et al., 2018).

Genome-wide transcriptomic profiling has contributed to our understanding of vitiligo pathogenesis. Previous studies revealed not only IFN-γ-induced gene expression and immune cell infiltration (Rashighi et al., 2014; Xu et al., 2022) but also melanocyte-intrinsic changes and WNT pathway dysregulation (Regazzetti et al., 2015). Keratinocyte remodeling is also found to be a factor (Singh et al., 2017). Meta-analysis of these transcriptomic profiles has the potential to identify consensus expression signatures of vitiligo. A previous rank-based meta-analysis of three microarray datasets identified candidate biomarkers (Zhang et al., 2021), but a broader comparison across platforms is needed to distinguish robust consensus signatures from study-specific signals.

We performed meta-analysis of six independent vitiligo transcriptomic studies (Table 1). Using Robust Rank Aggregation (Kolde et al., 2012), we identified consensus differentially expressed genes (DEGs) across diverse data types, characterized dysregulated pathways, and leveraged single-cell RNA-seq data from three studies (Shiu et al., 2022; Brunner et al., 2025; Xu et al., 2022) to validate signatures at the cell level. Our findings reveal that melanocyte loss is robustly detected across platforms while immune activation is study dependent.

**Table 1.** Studies Included in the Meta-Analysis.

| Study | Accession | Disease | Platform | Samples | Genes | DEGs<sup>a</sup> |
| --- | --- | --- | --- | --- | --- | --- |
| Brunner 2025 | GSE298871 | NSV | scRNA-seq | 5 pairs | 16,133 | 2 |
| Brunner 2025 (bulk) | Supp. Table | NSV | RNA-seq | 15 pairs | 19,773 | 1,344 |
| Singh 2017 | GSE75819 | Stable NSV | Illumina | 15 pairs | 16,312 | 3,154 |
| Regazzetti 2015 | GSE65127 | Active NSV | Affymetrix | 10 pairs | 13,908 | 141 |
| Rashighi 2014 | GSE53146 | Active vitiligo (FFPE) | Illumina | 5 V + 5 C | 12,707 | 0 |
| Xu 2021 | OMIX691 | NSV | scRNA-seq | 10 V + 5 H | 13,080 | 1,239 |
| Shiu 2022<sup>b</sup> | GSE203262 | Stable vitiligo (blister) | scRNA-seq | 5 pairs | 11,995 | 133 |

<sup>a</sup>Number of differentially expressed genes (FDR < 0.05 and $|\log_2 FC| > 0.58$ [FC > 1.5]). <sup>b</sup>Excluded from the final meta-analysis based on leave-one-out validation. C, control; V, vitiligo; H, healthy; NSV, nonsegmental vitiligo; scRNA-seq, single-cell RNA sequencing; FFPE, formalin-fixed paraffin-embedded.

## Results

### Study characteristics and data harmonization

We compiled transcriptomic data from seven independent vitiligo datasets comprising 125 samples across microarray, RNA-seq, and scRNA-seq (single-cell RNA-Seq) platforms (Table 1, Figure 1). Five datasets used paired designs comparing lesional and non-lesional skin within patients, while two used unpaired designs comparing skin samples from vitiligo patients with those from healthy controls.

**Fig. 1.**
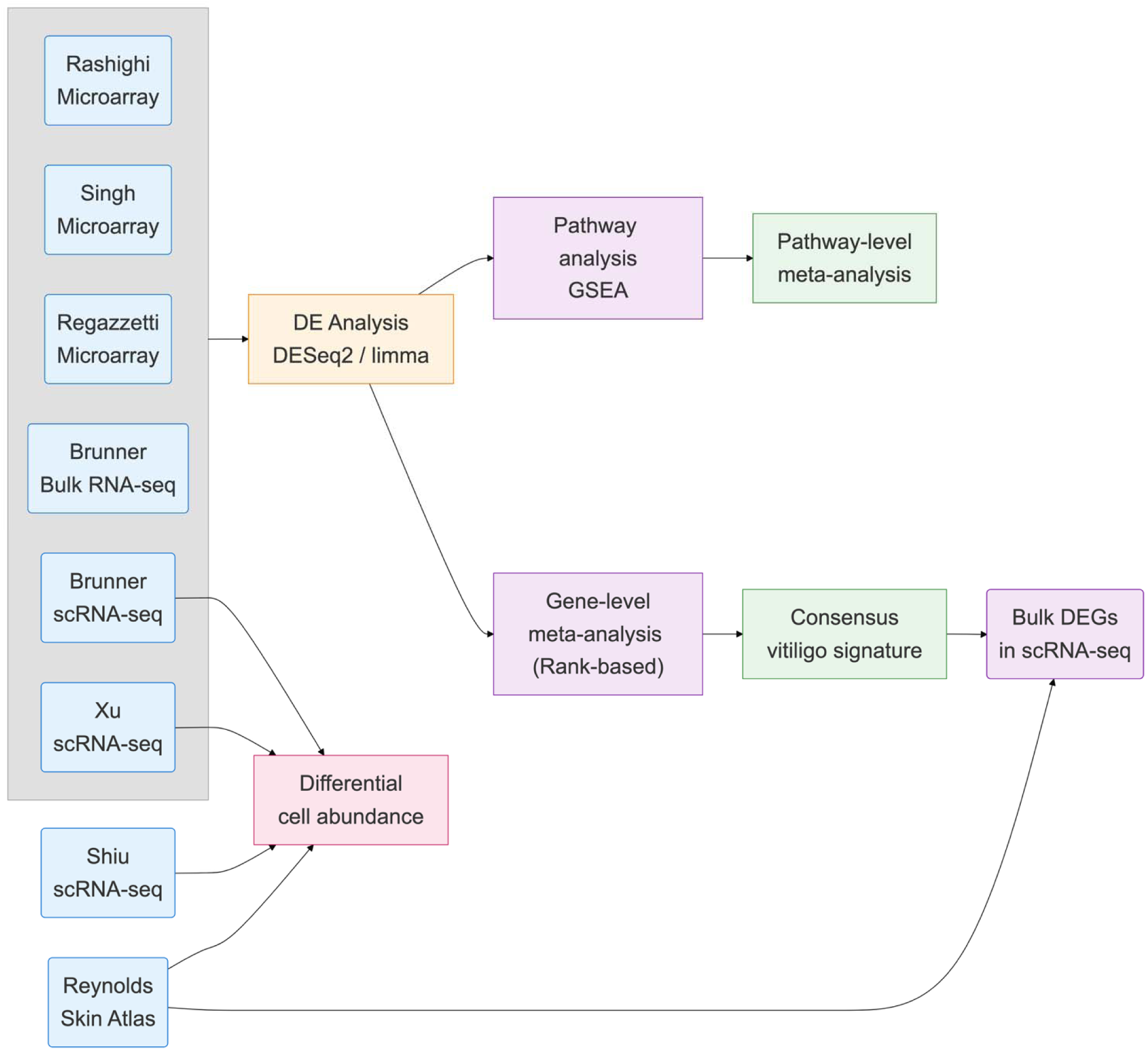
Meta-analysis workflow of transcriptomic studies of vitiligo skin.

Raw data were uniformly reprocessed using DESeq2 (Love et al., 2014) or limma (Ritchie et al., 2015), with stringent filtering of lowly expressed genes (Methods). For Brunner bulk RNA-seq data, we used the differential expression results from the original publication (Brunner et al., 2025) because count data are not publicly available. Results were harmonized to a common schema of gene symbols, log2 fold changes (FC), test statistics, and false discovery rates (FDRs), with 8,064 genes detected across all seven studies.

### Limited overlap in differentially expressed genes (DEGs)

Exploratory analysis of the harmonized results revealed substantial heterogeneity across studies. Statistical power varied considerably: studies with larger sample sizes showed strong enrichment for small p-values, while smaller studies exhibited near-uniform p-value distributions (Figure S1). Using conventional thresholds (FDR < 0.05, FC > 1.5), DEG counts ranged from 0 (Rashighi study) to 3,154 (Singh study), with limited overlap across studies, especially on upregulated genes (Table 1, Figures S2–S3). Pairwise log FC correlations were weak (−0.14 to 0.39; Figure S4), yet top-ranked genes showed consistent positioning across studies (Figures S5–S6). This pattern motivated rank-based rather than effect-size-based meta-analysis.

### Rank-based meta-analysis reveals direction-dependent robustness

For meta-analysis, genes were ranked by test statistic within each study, and Robust Rank Aggregation (RRA; Kolde et al., 2012) was used to identify genes ranking consistently higher (or lower) than expected by chance. RRA applied to 13,521 genes present in at least four of the seven studies identified 222 DEGs at FDR < 0.05: 140 downregulated genes, dominated by melanocyte markers, and 82 upregulated genes, enriched for interferon-stimulated genes.

To evaluate the robustness of the results, we systematically eliminated one dataset and repeated the meta-analysis. This leave-one-out (LOO) analysis revealed direction-dependent stability (Figure S7). Downregulated genes were more robust than upregulated genes. Removing the Shiu study led to a dramatic loss of upregulated genes (Jaccard = 0.07; only 6 of 82 retained), whereas downregulated genes remained intact (Jaccard = 0.58).

Investigation revealed that Shiu-specific upregulated genes were dominated by MHC class II molecules and T cell markers. This may be related to the use of suction blister sampling, which captures epidermal immune cells more efficiently than conventional biopsies. To obtain a consensus signature comparable across platforms, we excluded the Shiu dataset from the final meta-analysis.

### Downregulation of pigmentation genes is consistent

After excluding the Shiu dataset, RRA was applied to the remaining six studies, which identified 108 downregulated and 6 upregulated genes (Figure 2), revealing a fundamental asymmetry in the consensus vitiligo signature. See supplementary Table S1 for detailed results.

**Figure 2.**
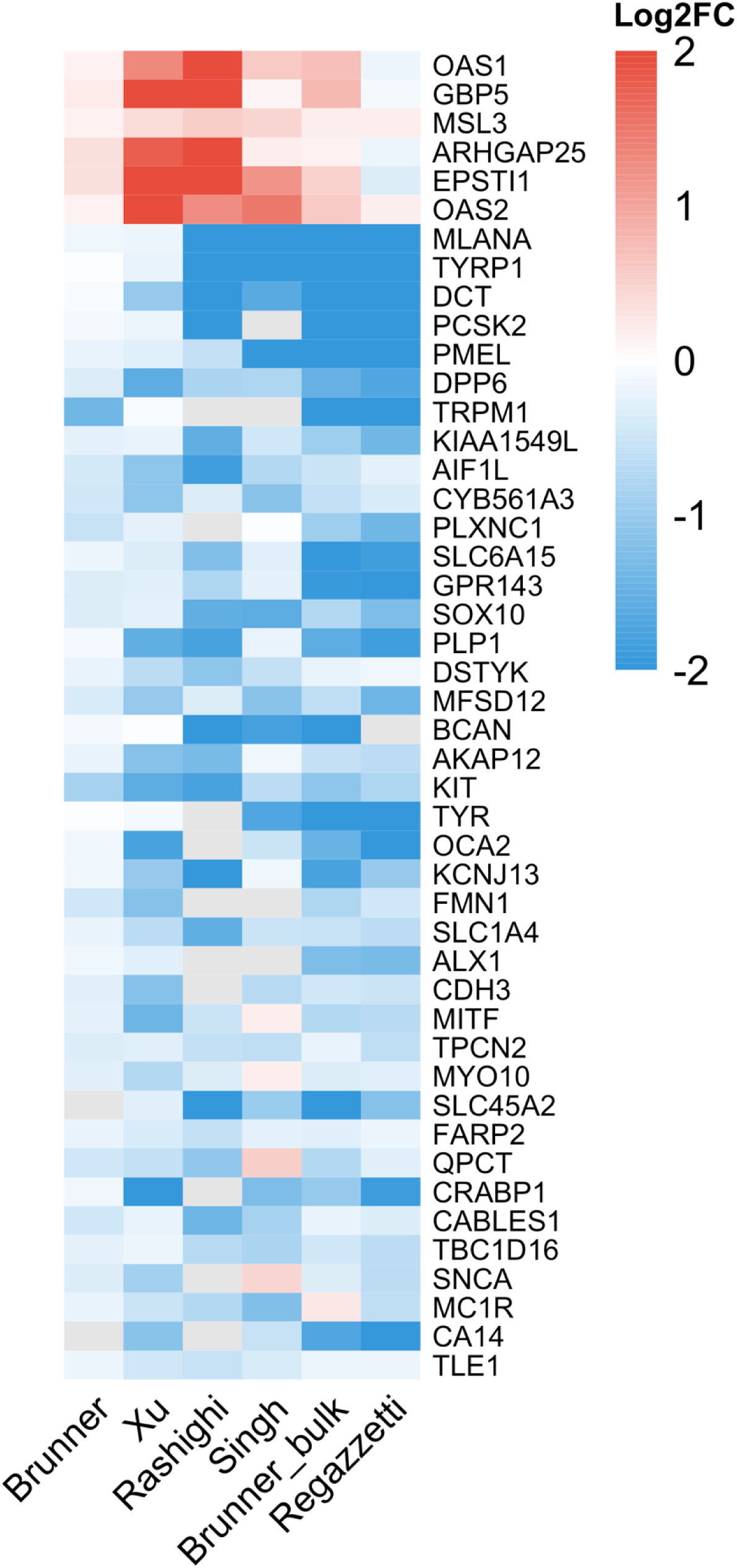
Cross-study log2 fold change heatmap for 6 significantly upregulated genes and the top 40 downregulated genes.

The top-downregulated genes include canonical melanocyte markers such as MLANA, TYRP1, DCT, PMEL, TRPM1, KIT, and TYR (Figure 2). These genes showed 100% direction concordance across all studies that detected them, with mean log2FC ranging from −1.6 to −2.7. Additional melanocyte genes (SOX10, MITF, OCA2, SLC45A2, GPR143) were also significantly downregulated. Over-representation analysis (ORA) of the 108 downregulated genes identifies Gene Ontology (GO) terms related to melanocyte biosynthesis (Figure S8). Unexpectedly, downregulated genes were also functionally enriched for terms like synaptic transmission and axon development.

In contrast, only six genes achieved significance in the upregulated direction: OAS1, OAS2, EPSTI1, GBP5, MSL3, and ARHGAP25 (Figure 2). These include interferon-stimulated genes (OAS1, OAS2, EPSTI1, GBP5), consistent with the established role of type I/II interferon signaling in vitiligo (Harris et al., 2012; Frisoli et al., 2020). The small number of robust upregulated genes may reflect variable immune cell capture across tissue extraction methods and a weaker underlying signal from infiltrating immune cells compared to the near-complete loss of melanocyte transcripts.

### Downregulation of neural development pathways in vitiligo

To identify dysregulated pathways, we performed Gene Set Enrichment Analysis (GSEA) using all genes ranked by a statistic derived from the meta-analysis. We used pi-value (−log (p) × log FC, Xiao et al., 2014), which incorporates both effect size and statistical significance while encoding direction. The top significantly enriched GO Biological Process terms are shown in Figure 3A. Activated pathways were dominated by type I interferon signaling and the virus defense response. Suppressed pathways include pigmentation, melanin biosynthesis, and, to a lesser extent, nervous system-related processes.

**Figure 3A.**
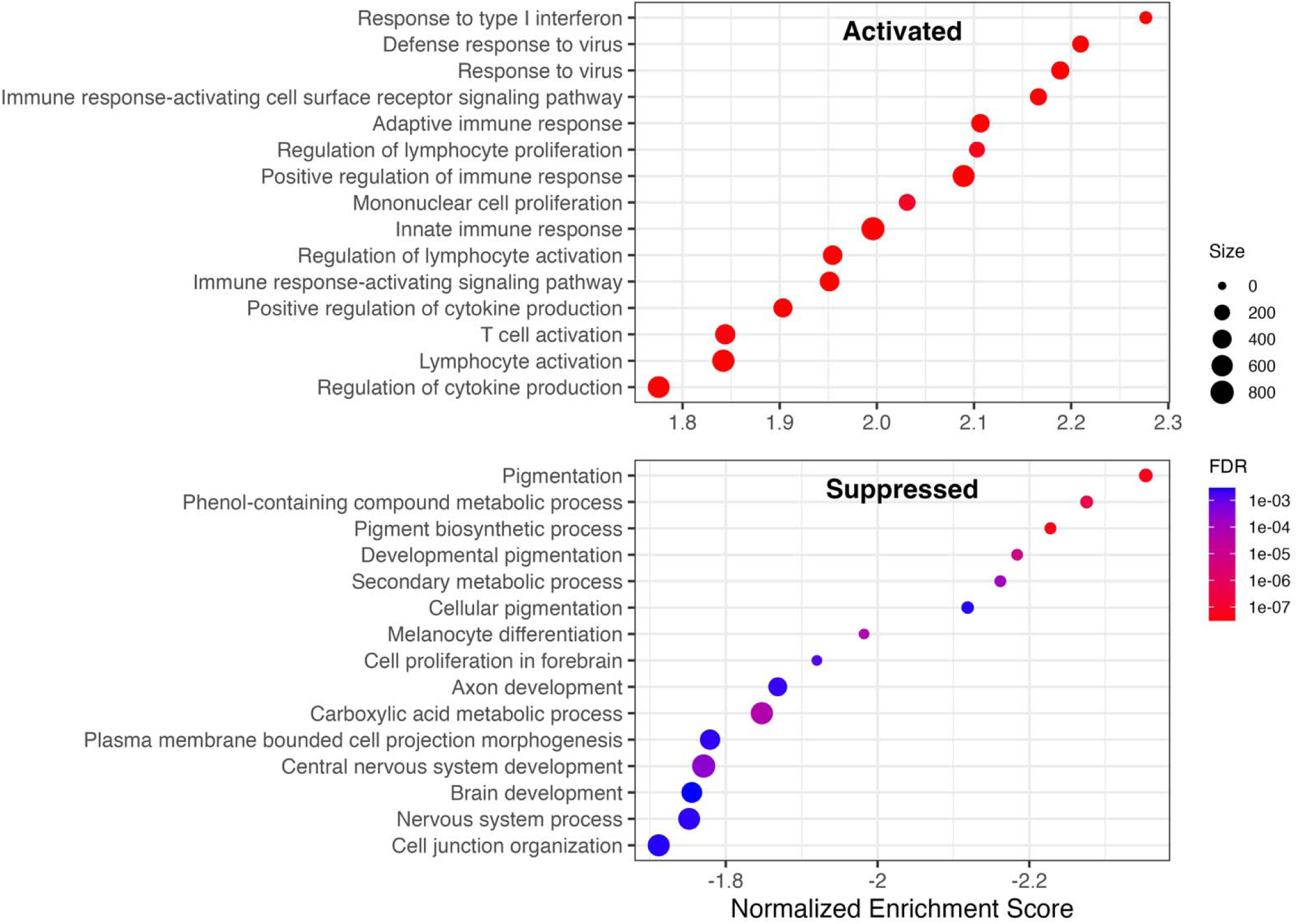
Gene Set Enrichment Analysis (GSEA) of genes ranked by meta-analysis. (A) Combined dot plot of GSEA results for GO Biological Process terms. The top panel shows activated pathways (positive normalized enrichment score; NES); the bottom panel shows suppressed pathways (negative NES).

**Figure 3B.**
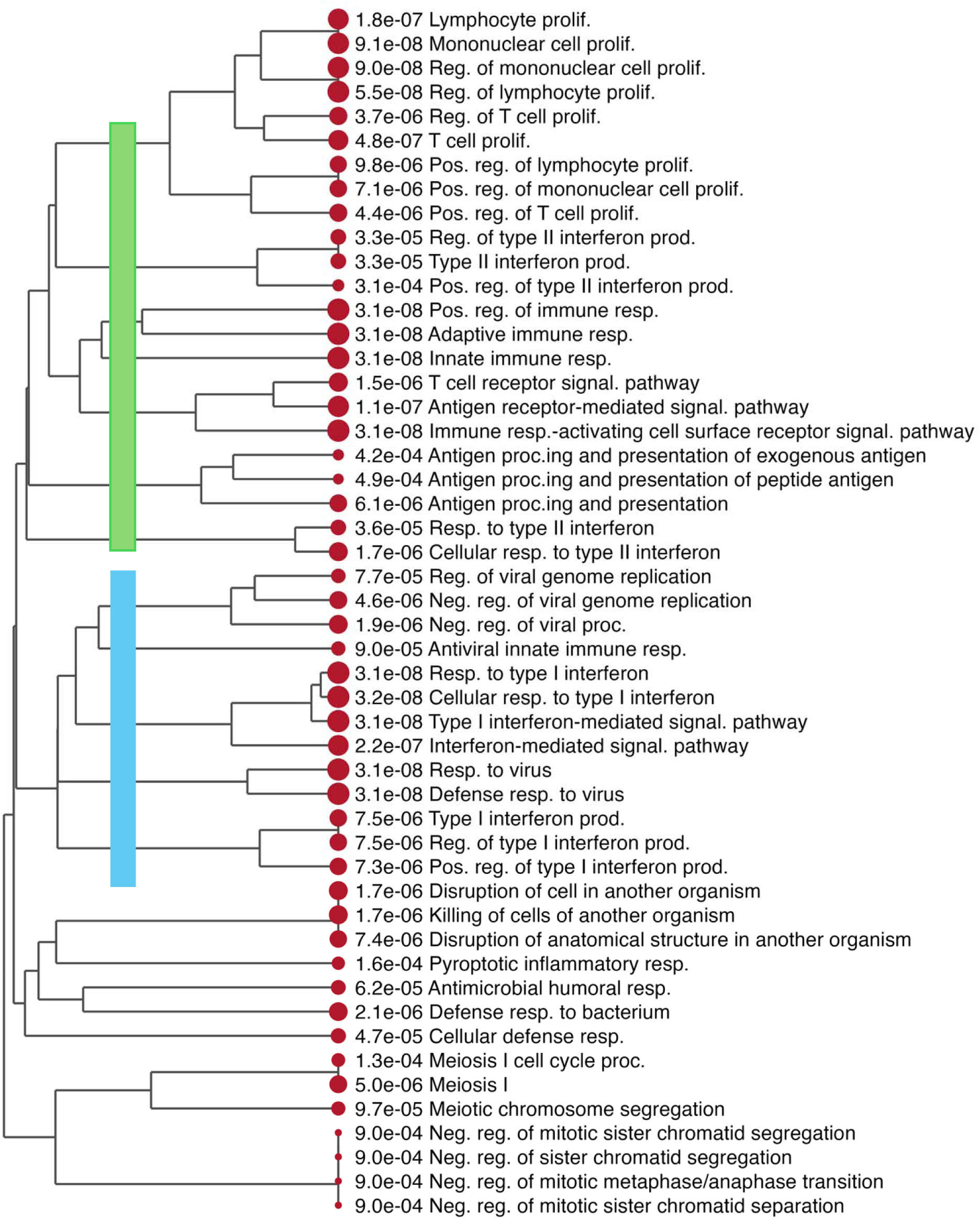
Gene Set Enrichment Analysis (GSEA) of genes ranked by meta-analysis. (B) Hierarchical tree visualization of activated GO terms, clustered by Jaccard similarity of gene membership. This forms two large clusters of terms for type I interferon and type II interferon responses, respectively.

**Figure 3C.**
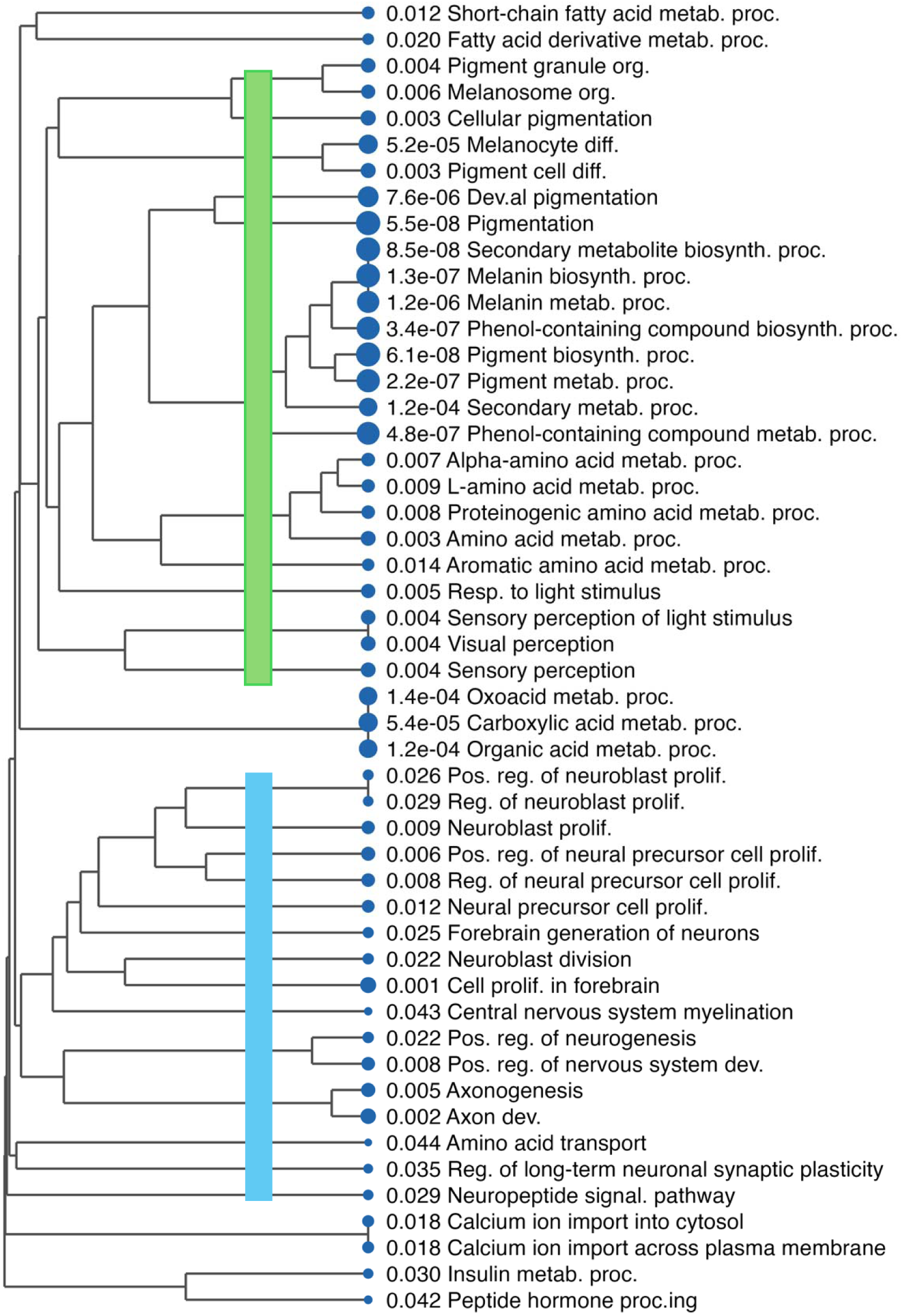
Gene Set Enrichment Analysis (GSEA) of genes ranked by meta-analysis. (C) Hierarchical tree visualization of suppressed GO terms. This shows that the neuronal-related terms form a cluster independent of the pigmentation cluster.

Hierarchical clustering of the top 50 enriched terms by gene overlap revealed distinct functional modules (Figure 3B–C). Activated terms clustered into type I and type II interferon pathways. Suppressed terms included a large melanocyte function cluster (pigmentation, melanin biosynthesis, melanosome organization) and, notably, a separate cluster of neural development pathways—axon development, neuron projection morphogenesis, and central nervous system development (Figure 3C). This unexpected finding could represent noise. However, leading-edge analysis confirmed minimal overlap: only 6 of 157 neural pathway genes (3.8%) were shared with melanocyte pathways. The neural-specific genes—including PLP1, NRXN3, and PLXNC1—showed significant downregulation (median log FC = −0.77, P < 4 × 10 ^5^) with 80% cross-study consistency, suggesting that the signal reflects genuine biology, albeit with smaller effect sizes than those of melanocyte genes. Because melanocytes derive from neural crest progenitors, the enrichment of neural-specific genes and pathways is likely driven by melanocyte loss. We demonstrated consistent downregulation of neuronal pathways in vitiligo driven by melanocyte loss.

### Pathway-level meta-analysis confirms cross-study concordance

To complement the gene-level analysis, we performed GSEA for each study individually and then conducted a meta-analysis of pathway rankings across studies. Of 5,126 Gene Ontology (GO) terms tested, 681 showed concordant enrichment (present in ≥4 studies, ≤1 discordant direction, ≥2 significant at FDR < 0.05). Per-study normalized enrichment scores (NES) of the top pathways are shown in Figure 4. Pigmentation pathways showed uniform suppression across all six studies, whereas immune pathways showed greater variability in concordance.

**Figure 4.**
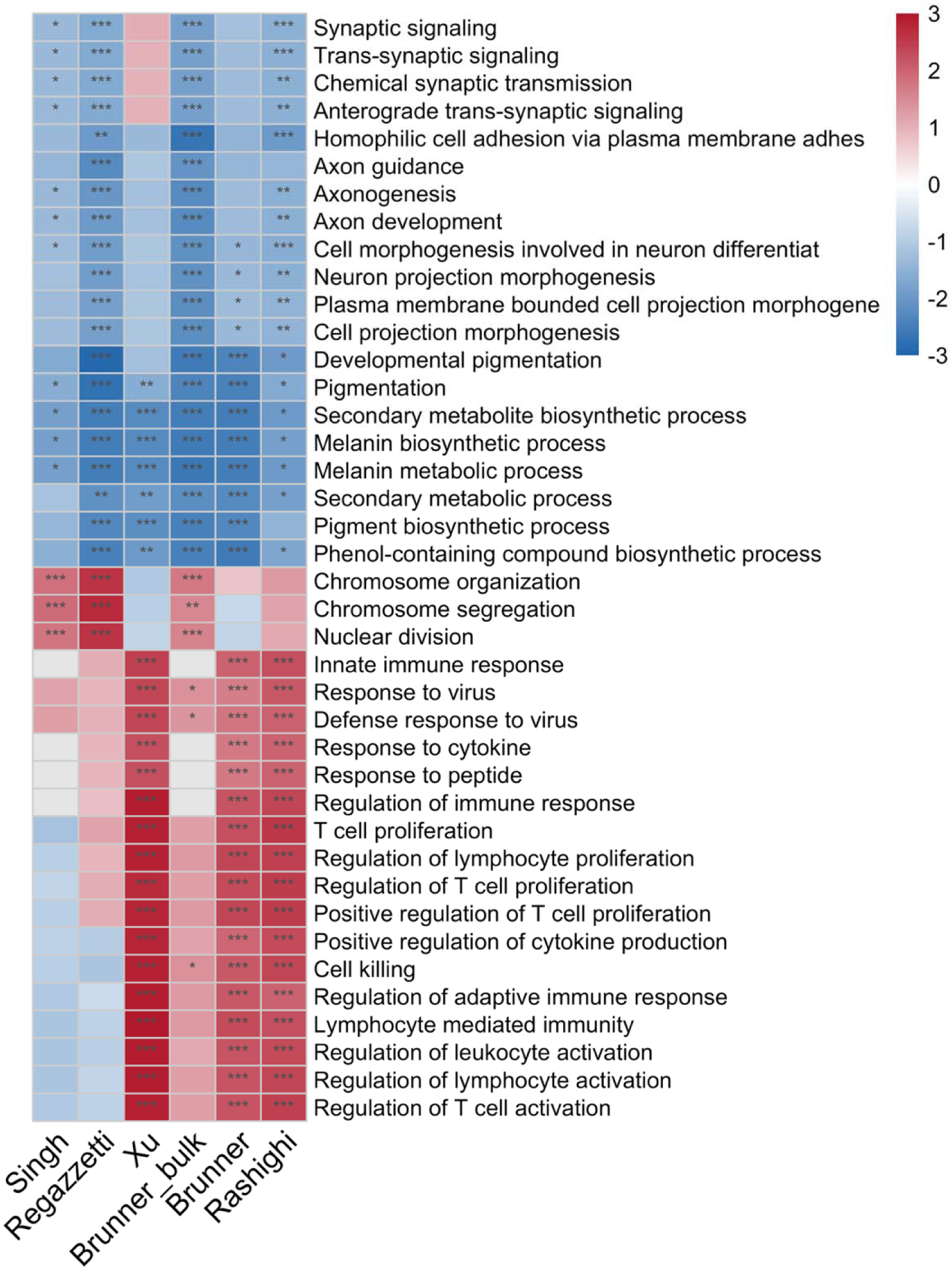
NES heatmap of the top 40 GO Biological Process terms from per-study GSEA analyses (20 activated, 20 suppressed) across six studies. Color indicates NES (red = activated, blue = suppressed); asterisks denote significance (*: FDR < 0.05, **: FDR < 0.01, ***: FDR < 0.001).

Per-study GAEA P-values were combined using Stouffer’s signed Z-method (Whitlock, 2005), where discordant effects cancel rather than accumulate significance. The top suppressed pathways were pigmentation (FDR = 4.9 × 10 ² ) and melanin metabolism (FDR = 1.0 × 10 ²¹); the top activated pathways were innate immune response (FDR = 1.7 × 10 ¹ ) and T cell proliferation (FDR = 3.3 × 10 ¹ ).

This analysis confirmed the unexpected suppression of neural development pathways in. vitiligo. Multiple neural pathways—axon development, axonogenesis, neuron projection morphogenesis—ranked among the most robustly suppressed, with effect sizes comparable to pigmentation. Critically, all six datasets showed independent suppression, with four to six reaching individual significance. Leading-edge analysis identified core contributors, including melanocyte markers (SOX10, DCT) alongside neural-associated genes (NGFR, L1CAM, PLP1, NRXN3, GPM6B).

### Melanocyte loss explains most downregulated genes

To investigate whether transcriptomic changes reflect altered cell-type composition, we analyzed cell-type abundance across three scRNA-seq datasets (Brunner, Shiu, Xu). Cells were annotated using reference-based label transfer from the Reynolds healthy skin atlas (Reynolds et al., 2021), and abundance changes between lesional and normal skin were tested (Figure 5A). Per-study p-values were combined using Stouffer’s signed Z-method (Whitlock, 2005).

As expected, melanocytes showed consistent depletion across all three studies (mean log2FC = - 2.87; combined FDR = 0.003), aligning with the transcriptomic signature dominated by downregulated melanocyte markers. Several immune cell types were significantly enriched, including regulatory T cells (Tregs, FDR = 0.007), cytotoxic T cells (FDR = 0.015), migratory dendritic cells (FDR = 0.015), and DC2 (FDR = 0.030), confirming the dual hallmarks of vitiligo: melanocyte loss accompanied by infiltration of antigen-presenting and cytotoxic immune cells.

To test whether melanocyte loss fully explains the downregulated signature, we examined the cell-type specificity of the 108 downregulated genes using percent detection across cell types in the healthy skin atlas (Figure 5B). The vast majority were detected far more frequently in melanocytes than in other cell types. This is confirmed by average expression analysis (Figure S10). Our analysis provides strong evidence that melanocyte loss drives the downregulated transcriptomic signature of vitiligo.

**Figure 5A.**
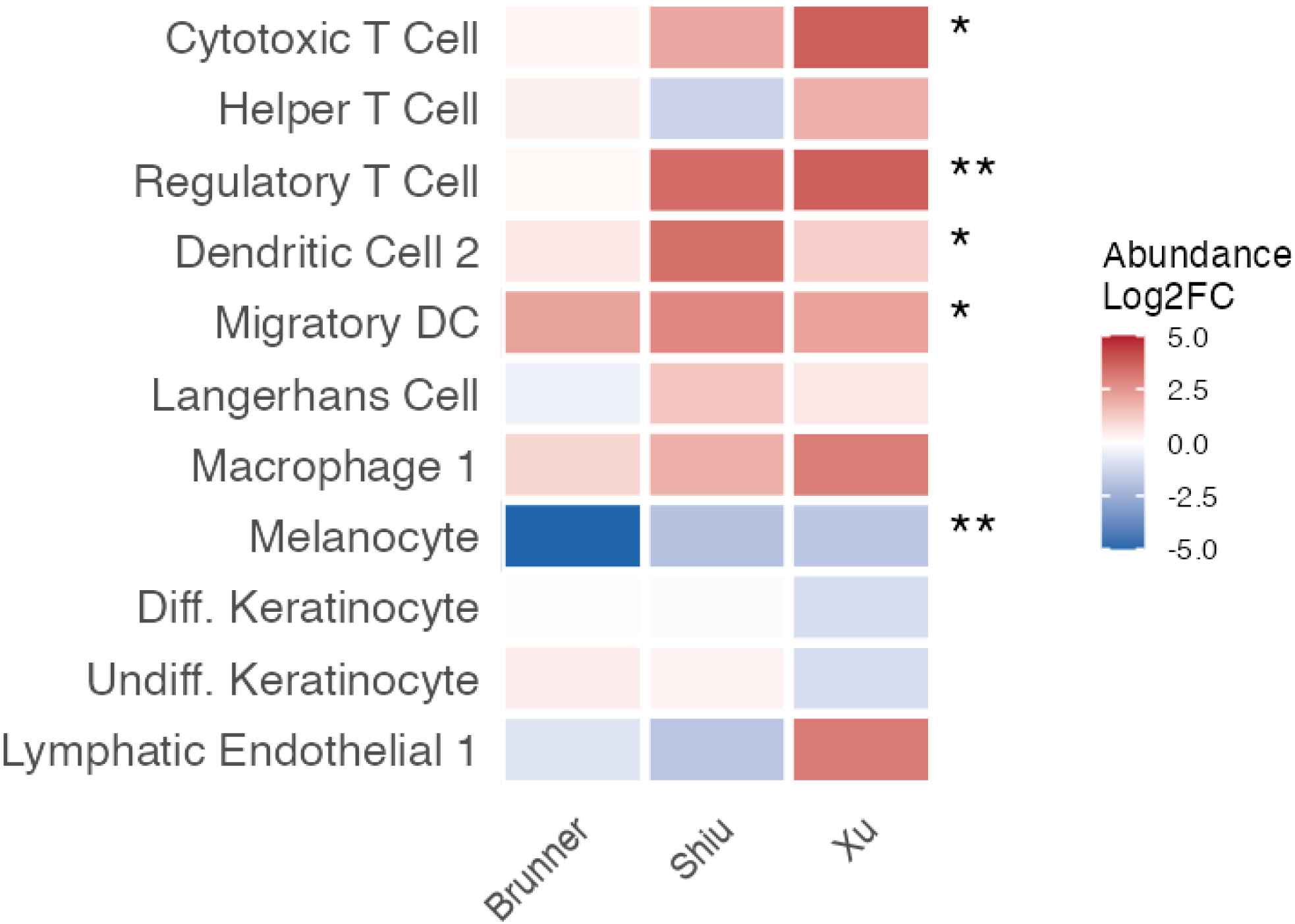
Changes in cell-type composition in vitiligo. (A) Heatmap showing log2 fold change in cell-type abundance between lesional and control skin across three scRNA-seq studies (Brunner, Shiu, Xu).

**Figure 5B.**
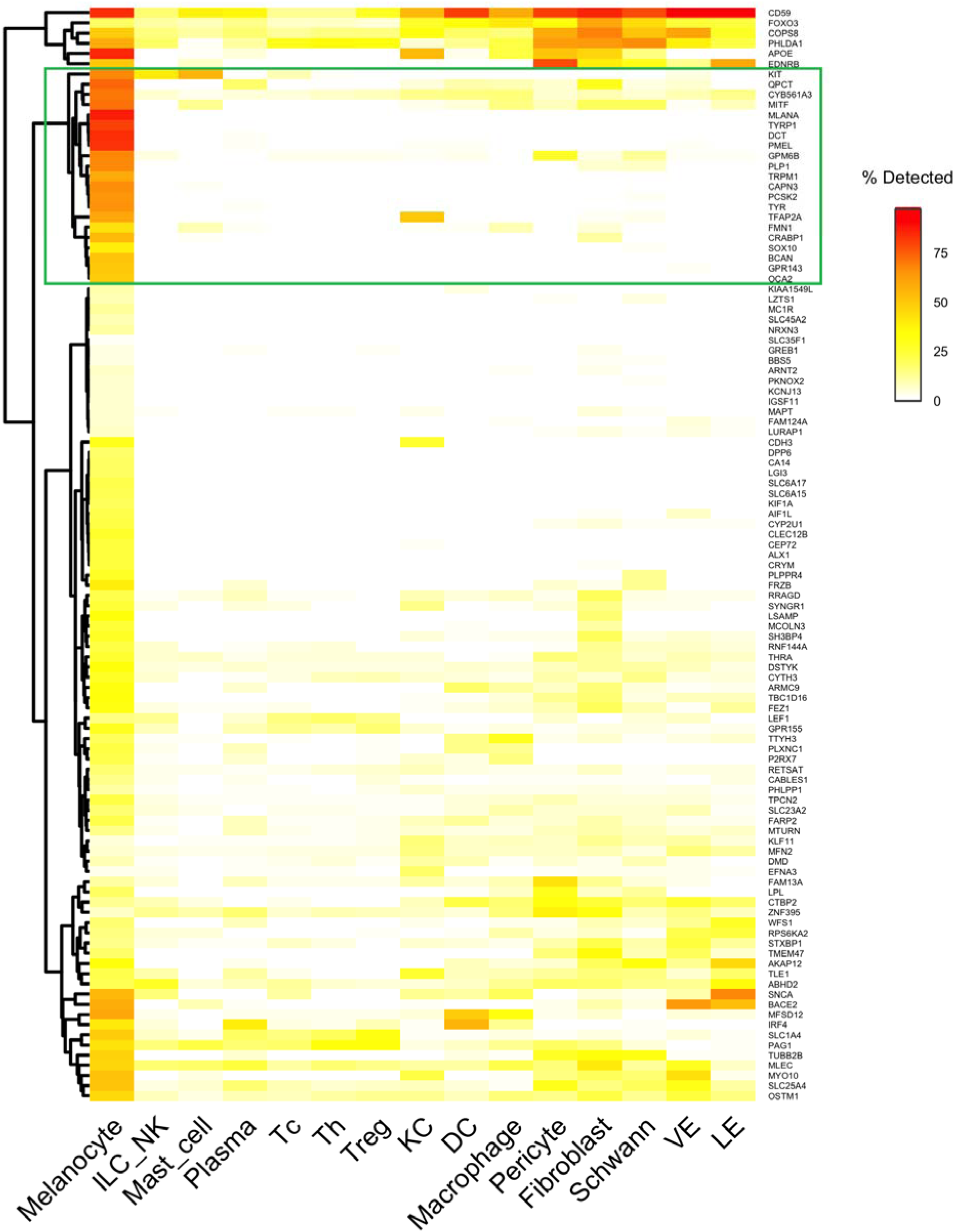
Changes in cell-type composition in vitiligo. (B) For the 108 genes downregulated in vitiligo, this heatmap shows detection rates across cell types. We identify a melanocyte-specific cluster containing 21 known and potentially novel marker genes.

### Downregulated genes include melanocyte markers

The heatmap of the detection rate of downregulated genes across cell types (Figure 5B) includes a distinct cluster of 21 genes with near-exclusive melanocyte expression (Table S2). This cluster included established pigmentation genes essential for melanin synthesis (TYR, TYRP1, DCT), melanosome structure (PMEL, TRPM1), and melanocyte transcription (MITF, SOX10, TFAP2A) (Kenny et al., 2022). Additional melanocyte-associated genes included the receptor tyrosine kinase KIT, melanosomal membrane proteins OCA2 and GPR143 (mutations in which cause oculocutaneous and ocular albinism, respectively), and the retinoic acid-binding protein CRABP1 (Collins and Watt, 2008). This 21-gene cluster, defined by near-exclusive melanocyte expression and consistent downregulation in vitiligo, is enriched for melanocyte markers.

Several genes in this cluster have documented neural functions yet have shown striking melanocyte specificity over Schwann cells—another neural crest-derived cell type in skin. BCAN (brevican, detected in 49.5% of melanocytes vs. 0.4% of Schwann cells), PCSK2 (63% vs. 1.1%), and PLP1 (66% vs. 6.7%) all showed >10-fold enrichment in melanocytes despite their canonical neural associations.

The cluster also contained less-characterized genes with emerging evidence of melanocyte expression. CYB561A3, a lysosomal ferrireductase recently identified as a pigmentation regulator through genome-wide association studies (GWAS) in African populations (Feng et al., 2024), was detected in 69% of melanocytes versus 8% of other cell types. QPCT (glutaminyl cyclase) showed 72% detection in melanocytes versus <10% in other cell types, a pattern validated across in the Reynolds atlas and three vitiligo scRNA-seq studies. QPCT protein has been detected in hyperpigmented skin (Yin et al., 2015) and is highly expressed in melanoma (Gillis, 2006), yet its function in normal melanocyte biology remains unexplored. These and other genes in the 21-gene cluster warrant functional investigation in the context of vitiligo.

## Discussion

The striking asymmetry of the vitiligo transcriptome—108 downregulated versus 6 upregulated genes—likely reflects a difference in signal characteristics. Melanocyte loss produces large, consistent transcriptomic changes, while immune-related changes appear weaker and more diverse.

Several factors may explain why immune signals are inconsistent. Statistical power is reduced by small sample sizes (5–15 patients) and a heterogeneous patient population (sex, age, health conditions). In paired designs, non-lesional skin is not immunologically quiescent, as resident memory T cells and subclinical immune networks persist in clinically normal-appearing skin (Gellatly et al., 2021; Migayron et al., 2025), compressing the lesional-versus-non-lesional differential. Uncontrolled variation in disease activity (Speeckaert et al., 2023) and anatomical site (Marella et al., 2025) further obscures immune signals. These observations do not diminish the established role of CXCR3 chemokine signaling in vitiligo (Speeckaert et al., 2023), but suggest that bulk transcriptomics with small cohorts may be insufficient to reliably capture immune signatures in this disease.

Our meta-analysis also identified a consistent suppression of neural development pathways. Re-analysis of single-cell datasets indicates that this signal can be largely explained by melanocyte expression profiles: genes driving the enrichment—PLP1, GPM6B, BCAN, PCSK2—were detected in >50% of melanocytes but <7% of Schwann cells (Figure 5B), consistent with melanocytes’ neural crest origin (Adameyko et al., 2009; Brombin and Patton, 2024). The roles of these genes in melanocyte biology warrant investigation.

This study has several limitations. The low cross-study effect size correlation (mean r = 0.11) justified rank-based meta-analysis, but this approach cannot provide pooled effect size estimates or distinguish weak consistent signals from strong signals in few studies. The heterogeneous comparisons—lesional versus non-lesional in paired studies, vitiligo versus healthy controls in unpaired studies—may capture different aspects of pathology. Platform differences, including pseudobulk aggregation and FFPE tissue, may introduce biases not fully captured by ranking.

In summary, meta-analysis across six heterogeneous transcriptomic studies reveals that the downregulated genes are almost entirely melanocyte-derived, including neural crest-associated genes whose expression in melanocytes has been underappreciated. Immune signatures, though detectable at the pathway level, are weak and variable at the gene level, likely reflecting differences in sampling method, disease activity, and study design. Integrating bulk meta-analysis with single-cell data proved essential for interpreting these signals and identified candidate melanocyte biomarkers—including CYB561A3, QPCT, and PCSK2—that warrant functional characterization. Future studies with larger, well-characterized cohorts and standardized sampling are needed to resolve the immune component of the vitiligo transcriptome.

## Materials and Methods

### Study Selection and Data Sources

We searched GEO and published literature for transcriptomic studies comparing vitiligo-affected skin to unaffected skin. Inclusion criteria were: (1) human samples, (2) genome-wide expression profiling (microarray or RNA-seq), (3) comparison of lesional versus non-lesional or vitiligo versus healthy control skin, and (4) availability of raw or processed expression data. Seven studies were initially included; one (Shiu) was subsequently excluded based on leave-one-out validation, yielding six studies comprising 115 samples (Table 1). Data were obtained from GEO (GSE298871, GSE75819, GSE65127, GSE53146, GSE203262) and the China National Center for Bioinformation (OMIX691). Platforms spanned Illumina BeadChip, Affymetrix microarray, bulk RNA-seq, and single-cell RNA-seq.

### Individual Study Re-Analysis

Five paired studies compared lesional to non-lesional skin within the same patients: Singh (n = 30, Illumina BeadChip, limma), Regazzetti (n = 20 from 10 pairs, Affymetrix, limma), Brunner bulk RNA-seq (n = 30, published DE results), Brunner pseudobulk (n = 10, DESeq2), and Shiu pseudobulk (n = 10 from 5 pairs after 1 patient excluded, DESeq2). Two unpaired studies compared vitiligo to control skin: Rashighi (n = 10, Illumina DASL for FFPE tissue, limma; 1 outlier excluded, 9 analyzed) and Xu (n = 15, pseudobulk DESeq2). For scRNA-seq datasets, pseudobulk profiles were generated by aggregating counts across all QC-passed cells within each sample. Microarray data were processed from raw files with standard normalization (quantile for Illumina, RMA for Affymetrix). For Brunner bulk RNA-seq, log2 fold changes and FDR-adjusted p-values were obtained from the original publication, as raw data were not available; test statistics were recovered from p-values using the inverse normal cumulative distribution function. Detailed processing for each study is described in Supplementary Methods.

### Data Harmonization and Meta-Analysis

Differential expression results were harmonized to a common schema: gene symbol, log2 fold change, standard error, test statistic (t-statistic for limma, Wald statistic for DESeq2), and p-values. Traditional inverse-variance meta-analysis was not appropriate because standard error miscalibration across platforms and low cross-study effect-size correlations (mean Pearson r = 0.11) were present. We therefore applied Robust Rank Aggregation (RRA), a non-parametric method that identifies genes ranked highly across studies without requiring effect-size standardization (Kolde et al., 2012). Genes were ranked within each study by absolute test statistic, and RRA was run separately for upregulated and downregulated genes; in each direction, genes with concordant fold change were placed at the top of the ranked list. Only genes present in at least four studies were included. P-values were corrected using the Benjamini-Hochberg method.

### Robustness Analysis

Leave-one-out analysis assessed stability by removing each study in turn and computing Jaccard similarity between the resulting gene set and the full analysis. The Shiu study was excluded from the final meta-analysis due to low Jaccard similarity (0.073 for upregulated genes), indicating that it disproportionately influenced the results.

### Pathway Enrichment Analysis

Gene Set Enrichment Analysis (GSEA) used the full ranked gene list (ordered by pi-value = median log2FC × −log10(RRA p-value); Xiao et al., 2014) to test Gene Ontology Biological Process and KEGG pathway enrichment using clusterProfiler (Wu et al., 2021). Over-representation analysis (ORA) tested meta-analysis significant genes (FDR < 0.05) against the same databases. GO terms were clustered by semantic similarity using simplifyEnrichment (Gu and Hübschmann, 2023). To assess whether suppressed neural development pathways reflected genuine biology, we performed leading-edge analysis. Genes from GSEA core enrichment were extracted for neural-related pathways (identified by keywords: neuro, axon, neural, nerve, synap, glia, brain, nervous) and melanocyte pathways (melano, pigment, melanin, tyrosin), and overlap was quantified using Jaccard similarity and a hypergeometric test. Effect sizes were compared across gene categories using Kruskal-Wallis and pairwise Wilcoxon tests.

### Pathway-Level Meta-Analysis

To assess pathway concordance across studies, GSEA was performed on each study individually. Per-study normalized enrichment scores were combined using Stouffer’s Z-method with signed Z-scores (Whitlock, 2005), ensuring discordant directions cancel rather than accumulate significance. Pathways were considered concordant if present in ≥4 studies with ≤1 discordant direction and significant in ≥2 studies.

### Cell Type Proportion Analysis from scRNA-seq data

To assess whether bulk transcriptomic signatures reflected changes in cell-type composition, we analyzed three scRNA-seq datasets (Xu, Brunner, Shiu). Cell types were annotated via reference-based label transfer from the Reynolds healthy skin atlas (Reynolds et al., 2021) using Seurat v5, with annotation confidence assessed using prediction scores and entropy filtering. Annotations were validated against canonical markers. Cell type proportions were compared between conditions using limma (paired studies) or propeller (Phipson et al., 2022; unpaired), with arcsine-square root transformation to stabilize variance. Cross-study evidence was combined using Stouffer’s Z-method. Details are provided in Supplementary Methods.

### Cell Type Specificity of Meta-Analysis Genes

To assess whether the downregulated meta-analysis signature reflects melanocyte loss, we examined the cell-type specificity of the 108 downregulated genes using percent detection (the percentage of cells with non-zero expression) and average expression across cell types from the Reynolds healthy skin atlas. Genes were hierarchically clustered (Ward’s D2 method), revealing a 21-gene cluster with near-exclusive melanocyte expression (Table S1).

### Statistical Analysis

All analyses were performed in R (v4.5.2). Multiple testing correction used the Benjamini-Hochberg method, with FDR < 0.05 considered significant. Key packages: limma (v3.66.0; Ritchie et al., 2015), DESeq2 (v1.48.0; Love et al., 2014), RobustRankAggreg (v1.2.1; Kolde et al., 2012), clusterProfiler (v4.18.4; Wu et al., 2021), fgsea (v1.36.2; Korotkevich et al., 2021), simplifyEnrichment (v1.10.0; Gu and Hübschmann, 2023), Seurat (v5.4.0; Hao et al., 2024), and speckle (v1.4.0; Phipson et al., 2022).

## Ethics Statement

This study is a secondary analysis of publicly available, de-identified datasets. No new human samples were collected.

## Data Availability Statement

This study is a secondary analysis of publicly available datasets. All source data are available from the following repositories:

- Gene Expression Omnibus (GEO): GSE298871 (Brunner pseudobulk), GSE75819 (Singh), GSE65127 (Regazzetti), GSE53146 (Rashighi), and GSE203262 (Shiu).
- China National Center for Bioinformation: OMIX691 (Xu).

Brunner bulk RNA-seq differential expression results were obtained from Table S3 of the original publication (Brunner et al., 2025). Read counts data were not publicly available at the time of analysis.

All analysis code is available at https://github.com/gexijin/vitiligo. The complete research process is tracked in the commit history. A Docker virtual machine image preserving the exact computing environment is available at https://hub.docker.com/repository/docker/gexijin/vitiligo/ to ensure reproducibility.

## Data Availability

No original data.

https://github.com/gexijin/vitiligo

## Acknowledgments

This work was supported by NIH grants (P20GM135008, R01HG013534, R41HG014101, and R43GM153076). Microsoft for Startups provided AI credits. The funders had no role in study design, data collection and analysis, decision to publish, or preparation of the manuscript.

## Author Contributions

**Xijin Ge:** Conceptualization, Methodology, Software, Formal Analysis, Data Curation, Writing – Original Draft, Writing – Review & Editing, Visualization, Funding Acquisition.

## Declaration of Generative AI Use

Generative AI was used extensively throughout this work for planning, coding, and writing. The following tools were used:

1. **Claude Code** (Anthropic) — Used for designing analytical approaches, writing and debugging R scripts for data processing, differential expression analysis, meta-analysis, pathway enrichment, and visualization, as well as creating the initial draft of the manuscript.
2. **ChatGPT** (OpenAI) — Used to assist in planning the study design and code review (Codex CLI).

All prompts, conversation logs, and custom slash commands developed to streamline recurring tasks are available in the GitHub repository. Analyses were conducted between December 2024 and February 2026. The author has reviewed and edited all AI-generated content and takes full responsibility for the accuracy and integrity of this work.

## Abbreviations

DE: differential expression
DEG: differentially expressed gene
FC: fold change
FDR: false discovery rate
FFPE: formalin-fixed paraffin-embedded
GEO: Gene Expression Omnibus
GO: Gene Ontology
GSEA: Gene Set Enrichment Analysis
GWAS: genome-wide association study
IFN: interferon
KEGG: Kyoto Encyclopedia of Genes and Genomes
LOO: leave-one-out
MHC: major histocompatibility complex
NES: normalized enrichment score
ORA: over-representation analysis
RMA: Robust Multi-array Average
RRA: Robust Rank Aggregation
scRNA-seq: single-cell RNA sequencing

