## Supplemental materials for "Melanocyte loss dominates the vitiligo transcriptome: a rank-based meta-analysis"

Xijin Ge

### Supplementary Materials

#### Study Selection and Data Sources

We searched the Gene Expression Omnibus (GEO) and published literature for transcriptomic studies comparing vitiligo-affected skin to unaffected skin. Inclusion criteria were: (1) human samples, (2) genome-wide expression profiling (microarray or RNA-seq), (3) comparison of lesional versus non-lesional skin or vitiligo versus healthy control skin, and (4) availability of raw or processed expression data. Seven studies meeting these criteria were initially included; one (Shiu) was subsequently excluded based on leave-one-out validation, yielding a final meta-analysis of six studies (Table 1).

Data were obtained from GEO (GSE298871, GSE75819, GSE65127, GSE53146, GSE203262) and the China National Center for Bioinformation (OMIX691). Platforms included Illumina HumanHT-12 BeadChip, Affymetrix Human Genome U133 Plus 2.0, bulk RNA-seq, and single-cell RNA-seq (10x Genomics).

#### Individual Study Processing

##### *Singh (GSE75819)*

Microarray expression data from Singh et al. (2017) was obtained from GEO, comprising 30 samples from 15 vitiligo patients with matched lesional and non-lesional skin biopsies profiled on the Illumina HumanHT-12 v3 BeadChip platform. Raw non-normalized intensity data were log2-transformed with offset (+1) and quantile-normalized across samples using limma. Of 48,803 probes, 29,254 (59.9%) passed expression filtering requiring normalized intensity above the global 40th percentile in at least 30% of samples. Differential expression analysis employed limma (Ritchie et al., 2015) with a paired design (~ patient + condition), followed by empirical Bayes moderation and Benjamini-Hochberg FDR correction. Probes were annotated using illuminaHumanv3.db and collapsed to gene level by selecting the probe with smallest p-value per gene, yielding 16,312 unique genes.

##### *Regazzetti (GSE65127)*

Microarray expression data from Regazzetti et al. (2015) was obtained from GEO, comprising 40 samples on the Affymetrix Human Genome U133 Plus 2.0 Array: 10 healthy volunteers and 10 vitiligo patients with lesional, perilesional, and non-lesional skin biopsies each. Quality control using NUSE and RLE metrics identified 2 outlier samples from healthy controls, which were excluded, yielding 38 samples. Raw CEL files were processed with RMA normalization including background correction, quantile normalization, and median polish summarization across 54,675 probesets. For the lesional versus non-lesional comparison, differential expression was performed using limma (Ritchie et al., 2015) with a paired design on 10 vitiligo patients. Probes were annotated using hgu133plus2.db and collapsed to gene level by selecting the probe with smallest p-value per gene, yielding 13,908 unique genes. Perilesional samples were available but not used in the meta-analysis.

##### *Rashighi (GSE53146)*

Microarray expression data from Rashighi et al. (2014) was obtained from GEO, comprising 10 formalin-fixed, paraffin-embedded (FFPE) skin samples profiled on the Illumina HumanHT-12 WG-DASL V4.0 platform designed for degraded RNA. Samples included 5 vitiligo lesional skin specimens with active inflammatory infiltrate and 5 age- and site-matched control skin specimens. One control sample was excluded as an outlier based on convergent evidence from multiple QC metrics (intensity, detection rate, PCA, and hierarchical clustering), leaving 9 samples for analysis. Expression data were log2-transformed with offset and quantile-normalized. Probes were filtered to retain those detected (detection $p < 0.05$) in at least 5 of 9 samples; no additional expression-level threshold was applied given the limited sample size. Differential expression used limma (Ritchie et al., 2015) with a cell means model contrasting vitiligo versus control. Probes were annotated using illuminaHumanv4.db and collapsed to gene level by selecting the probe with smallest p-value per gene, yielding 12,707 unique genes. No genes reached FDR < 0.05 significance due to limited statistical power, though nominally significant genes showed biologically coherent patterns and contributed to rank-based meta-analysis.

##### *Brunner Bulk RNA-seq*

Bulk RNA-seq differential expression results from Brunner et al. (2025) were obtained directly from the published study, comprising 30 samples from 15 patients with matched lesional and non-lesional skin biopsies. The original study performed differential expression using limma (Ritchie et al., 2015) and provided log2 fold changes, raw p-values, and FDR-adjusted p-values for 19,773 genes. Raw sequencing data were not publicly available. Test statistics were recovered from raw p-values using the inverse normal cumulative distribution function ($z = \text{qnorm}(1 - p/2)$), with sign determined by log2 fold change direction. Standard errors were estimated from log2 fold changes divided by recovered z-scores, with absolute z-scores floored at 0.1 to avoid extreme standard errors and standard errors capped at 10 to exclude unreliable estimates.

##### *Brunner Pseudobulk (GSE298871)*

Single-cell RNA sequencing data from Brunner et al. (2025) was obtained from GEO, comprising skin biopsies from 17 samples including healthy controls and vitiligo patients with lesional and non-lesional samples. For cell-level quality control, cells were filtered using lenient thresholds: minimum 200 genes detected, maximum 4,000 genes (to remove doublets), and maximum 25% mitochondrial reads. This filtering retained 21,889 of 24,554 cells (89.1%). For the meta-analysis, only the 5 patients with complete paired lesional/non-lesional data were included. Pseudobulk samples were generated by aggregating single-cell counts across all QC-passed cells within each patient-condition combination, yielding 10 pseudobulk samples. Gene filtering retained 16,133 genes with CPM ≥ 2 in ≥ 30% of samples and minimum 20 total counts. Differential expression was performed using DESeq2 (Love et al., 2014) with a paired design (~ patient_id + condition). Log2 fold change shrinkage was applied using the normal method.

##### *Shiu Pseudobulk (GSE203262)*

Single-cell RNA sequencing data from Shiu et al. (2022) was obtained from GEO, comprising matched lesional and non-lesional suction blister samples from six stable vitiligo patients. Cell-level quality control applied identical thresholds to other scRNA-seq studies: minimum 200 genes, maximum 4,000 genes, and maximum 25% mitochondrial reads, retaining 17,458 of 18,297 cells (95.4%). One patient (Patient G) was excluded as an outlier due to critically low library size (27,431 reads versus 0.7-28 million for other samples), yielding 10 pseudobulk samples from 5 patients. Gene filtering retained 11,995 genes with CPM ≥ 2 in ≥ 30% of samples. Differential expression was performed using DESeq2 with a paired design (~ patient + condition). Apeglm shrinkage (Zhu et al., 2019) was applied for log2 fold change estimation; however, maximum likelihood estimates (unshrunk) were used for meta-analysis harmonization. This study was subsequently excluded from the final meta-analysis based on leave-one-out validation.

##### *Xu Pseudobulk (OMIX691)*

Single-cell RNA sequencing data from Xu et al. (2022) was obtained from the China National Center for Bioinformation, comprising skin samples from 15 individuals (5 healthy controls, 10 vitiligo patients). Cell-level quality control applied the same thresholds as other scRNA-seq studies: minimum 200 genes, maximum 4,000 genes, and maximum 25% mitochondrial reads, retaining 50,920 of 52,845 cells (96.4%). Pseudobulk samples were generated by summing gene counts across all QC-passed cells within each sample, yielding 15 pseudobulk samples. Gene filtering retained 13,080 genes with CPM ≥ 2 in ≥ 30% of samples. Differential expression was performed using DESeq2 with an unpaired design (~ condition) comparing vitiligo versus healthy. Apeglm shrinkage (Zhu et al., 2019) was applied for log2 fold change estimation.

#### Data Harmonization

Differential expression results from all studies were harmonized to a common schema containing: gene symbol (uppercase HGNC), log2 fold change, standard error, test statistic (t-statistic for limma, Wald statistic for DESeq2), raw p-value, and FDR-adjusted p-value. For DESeq2 studies, the Wald test statistic and p-values are computed from the maximum likelihood estimate and are unaffected by LFC shrinkage; the primary meta-analysis ranking by test statistic is therefore independent of the shrinkage method used. For Brunner bulk, which provided only log2 fold changes and p-values from the original publication, test statistics were recovered as described in the individual study section above. Standard errors were derived from log2FC divided by test statistic, with extreme values capped.

#### Robust Rank Aggregation Meta-Analysis

Traditional inverse-variance weighted meta-analysis was not appropriate due to standard error miscalibration across platforms (limma SEs approximately 5-fold smaller than DESeq2 SEs for comparable genes) and low cross-study effect size correlations (mean Pearson $r = 0.11$). Instead, we applied Robust Rank Aggregation (RRA), a non-parametric method that identifies genes ranked consistently highly across studies without requiring effect size standardization (Kolde et al., 2012).

Genes were ranked within each study by absolute test statistic, with separate analyses for upregulated and downregulated directions. For each direction, genes with concordant direction were placed at the top of the ranked list; genes with opposite direction were appended at the bottom. RRA p-values were calculated using the aggregateRanks function from the RobustRankAggreg R package, with Benjamini-Hochberg correction for multiple testing. Only genes present in at least four studies were included.

#### Leave-One-Out Robustness Analysis

To assess stability, we performed leave-one-out (LOO) analysis, removing each study in turn and recalculating RRA on the remaining studies. Jaccard similarity was computed between each LOO gene set and the full analysis, separately for upregulated and downregulated genes. Studies whose removal caused substantial loss of significant genes (low Jaccard similarity) were flagged as potentially influential.

The Shiu study was identified as an outlier based on LOO analysis (Jaccard similarity = 0.073 for upregulated genes when removed) and was excluded from the final 6-study meta-analysis. Investigation of Shiu-specific genes was performed using over-representation analysis to characterize the biological pathways unique to this study.

#### Pathway Enrichment Analysis

Gene set enrichment was performed using two complementary approaches. Over-representation analysis (ORA) tested whether significant genes (FDR $< 0.05$ from RRA) were enriched for Gene Ontology Biological Process terms or KEGG pathways, using Fisher's exact test with all genes tested in RRA as background. Gene Set Enrichment Analysis (GSEA) used the full ranked gene list (ranked by pi-value = median log2FC across studies × -log10(RRA p-value); Xiao et al., 2014) to test for coordinated enrichment without requiring significance thresholds.

GSEA was performed using clusterProfiler (v4.18.4; Wu et al., 2021) with gene set size limits of 15–800; ORA used default clusterProfiler limits of 10–500. Multiple testing correction used the Benjamini-Hochberg method. For visualization, the top 100 suppressed GO BP terms (NES $< 0$, FDR $< 0.05$) were clustered by semantic similarity (Sim_Relevance_2006 measure) using binary cut clustering from simplifyEnrichment (Gu and Hübschmann, 2023).

To assess whether suppressed neural development pathways reflected genuine biology or annotation overlap with melanocyte pathways (due to shared neural crest origin), we performed leading edge analysis. Genes from GSEA core enrichment were extracted for neural-related pathways (identified by keywords: neuro, axon, neural, nerve, synap, glia, brain, nervous) and melanocyte pathways (melano, pigment, melanin, tyrosin). Overlap was quantified using Jaccard similarity and tested with a hypergeometric test. Effect sizes (median log2FC) were compared across gene categories using Kruskal-Wallis and pairwise Wilcoxon tests.

#### Pathway-Level Meta-Analysis

To assess pathway concordance independent of gene-level aggregation, GSEA was performed on each study individually using test statistic ranking for GO Biological Process terms and KEGG pathways (gene set size 15–800). Per-study normalized enrichment scores (NES) and p-values were assembled into concordance matrices. Pathways were considered concordant if present in ≥4 studies, showing ≤1 discordant direction, and significant (FDR < 0.05) in ≥2 studies.

Per-study results were combined using Stouffer's Z-method with signed Z-scores (Whitlock, 2005):

$$z_i = \text{sign}(\text{NES}_i) \times \Phi^{-1}(1 - p_i)$$

$$Z_{\text{combined}} = \frac{\sum z_i}{\sqrt{k}}$$

where $p_i$ is the raw GSEA p-value, $\text{NES}_i$ is the normalized enrichment score for study $i$, $\Phi^{-1}$ is the inverse standard normal CDF, and $k$ is the number of studies with valid data for that pathway. This approach ensures pathways with discordant directions across studies cancel out rather than accumulate significance. Combined p-values were converted from the combined Z-score using a two-tailed test and corrected for multiple testing using the Benjamini-Hochberg method.

#### Single-Cell RNA-seq Cell Type Proportion Analysis

To validate bulk transcriptomic findings at the cellular level and assess cell type composition changes in vitiligo, we analyzed three independent scRNA-seq datasets (Xu, Shiu, Brunner) using reference-based cell type annotation and compositional analysis.

##### *Reference Atlas*

The Reynolds healthy skin atlas (Reynolds et al., 2021) was used as reference for cell type annotation. This atlas comprises 195,739 cells from healthy adult skin with 34 annotated cell types spanning keratinocytes, immune cells (T cells, dendritic cells, macrophages, Langerhans cells), stromal cells (fibroblasts, pericytes, endothelial cells), and melanocytes. The atlas was converted from h5ad format to Seurat, log-normalized (scale factor 10,000), and processed to identify 3,000 highly variable genes for dimensionality reduction (50 PCA components).

##### *Reference-Based Label Transfer*

Cell type annotations were transferred from the Reynolds atlas to each vitiligo dataset using Seurat v5 (Hao et al., 2024) label transfer. For each query dataset, data were normalized, 3,000 variable features identified, and transfer anchors found using canonical correlation analysis (CCA) in PCA space (dims 1:50). Cell type labels and prediction scores were transferred from the reference.

Annotation confidence was assessed using a two-tier approach: (1) prediction score (maximum probability across cell types), and (2) prediction entropy (Shannon entropy of the full probability distribution). Cells were classified as reliable only if prediction score ≥ 0.5 AND entropy < 1.8 (approximately 60% of maximum possible entropy). This dual criterion identified 115 ambiguous cells (0.1% of total) across all datasets.

##### *Annotation Validation*

Transferred annotations were validated by testing enrichment of canonical markers in assigned cell types. For each cell type and canonical marker, we calculated fold enrichment (mean expression in annotated cells / mean expression in other cells) and significance (one-sided Wilcoxon rank-sum test). Validation criteria were: PASS (fold enrichment ≥ 1.5), WEAK (enrichment < 1.5), or FAILED (marker essentially absent: <5% expressing AND mean expression < 0.01).

Per-cell marker filtering was additionally applied: cells expressing zero canonical markers for their assigned type were relabeled as "Ambiguous" and excluded from downstream analysis. This filtering identified misclassified cells, particularly in Brunner where 45% of cells initially labeled as melanocytes expressed zero melanocyte markers (MLANA, PMEL, TYR, DCT, MITF), consistent with melanocyte depletion in lesional samples.

Overall, 90% of marker-celltype validations passed, with failures concentrated in very small populations (n = 1-2 cells) where dropout is expected.

##### *Cell Type Proportion Statistical Analysis*

Cell type proportions were calculated per sample using high-confidence cells (prediction score ≥ 0.5). Statistical testing accounted for study design:

**Paired studies (Shiu, Brunner):** limma with design matrix `~ patient + group` to block by patient. Only patients with both conditions were included (Shiu: 5 pairs after excluding Patient G; Brunner: 5 pairs after excluding one patient with only lesional samples). Proportions were arcsin-square root transformed for variance stabilization, and empirical Bayes moderated t-tests provided robust inference with small samples.

**Unpaired study (Xu):** propeller (Phipson et al., 2022) from the speckle package was used for two-group comparison (10 vitiligo vs 5 healthy), employing the same arcsin-square root transformation and precision weighting based on sample size.

Log2 fold changes were calculated from raw proportions with a small pseudocount (0.01) to handle zero proportions. Multiple testing correction used the Benjamini-Hochberg method within each study.

##### *Cross-Study Meta-Analysis of Cell Type Proportions*

Evidence across studies was combined using Stouffer's signed Z-score method. Two-sided p-values from proportion tests were converted to z-scores using z = qnorm(p/2, lower.tail = FALSE) and signed by the direction of log2FC, then combined as Z = sum(z_i) / sqrt(k). Combined p-values were FDR-corrected across cell types.

#### Cell Type Specificity of Meta-Analysis Genes

To assess whether the downregulated meta-analysis signature reflects melanocyte loss, we examined cell type specificity of the 108 downregulated genes (FDR < 0.05) using the Reynolds healthy skin atlas. Fine cell type annotations from the atlas were grouped into major categories (e.g., keratinocyte subtypes combined as KC, dendritic cell subtypes combined as DC) to reduce sparsity.

For each gene and cell type, percent detection was calculated as the percentage of cells with non-zero counts in the raw count matrix. Average expression was calculated as the mean of log-normalized expression values across cells of each type. Both metrics were assembled into gene × cell type matrices and visualized as heatmaps using pheatmap with hierarchical clustering (Ward's D2 method, Euclidean distance) on both rows (genes) and columns (cell types).

A cluster of 21 genes with near-exclusive melanocyte expression was identified from the dendrogram of the percent detection heatmap. These genes showed high percent detection in melanocytes (38–86%) compared to other cell types (0.04–8%), confirming melanocyte loss as the primary driver of the downregulated signature (Table S1).

#### Statistical Analysis

All statistical analyses were performed in R (v4.5.2). Multiple testing correction used the Benjamini-Hochberg method with FDR < 0.05 considered significant unless otherwise noted. Key packages included: limma (v3.66.0; Ritchie et al., 2015) for microarray analysis and cell type proportion testing, DESeq2 (v1.48.0; Love et al., 2014) for RNA-seq analysis, RobustRankAggreg (v1.2.1; Kolde et al., 2012) for rank aggregation, clusterProfiler (v4.18.4; Wu et al., 2021) for pathway enrichment, fgsea (v1.36.2; Korotkevich et al., 2021) for GSEA, simplifyEnrichment (v1.10.0; Gu and Hübschmann, 2023) for GO term clustering, Seurat (v5.4.0; Hao et al., 2024) for single-cell analysis and label transfer, and speckle (v1.4.0; Phipson et al., 2022) for cell type proportion testing with propeller.

#### Supplementary Tables

##### Table S1

**Title:** Complete Meta-Analysis Results for All Genes

##### Table S2

Expression characteristics of the 21-gene melanocyte marker cluster identified by hierarchical clustering of RRA-downregulated genes.

### Supplementary Figures

#### Figure S1. Distribution of Differential Expression Statistics

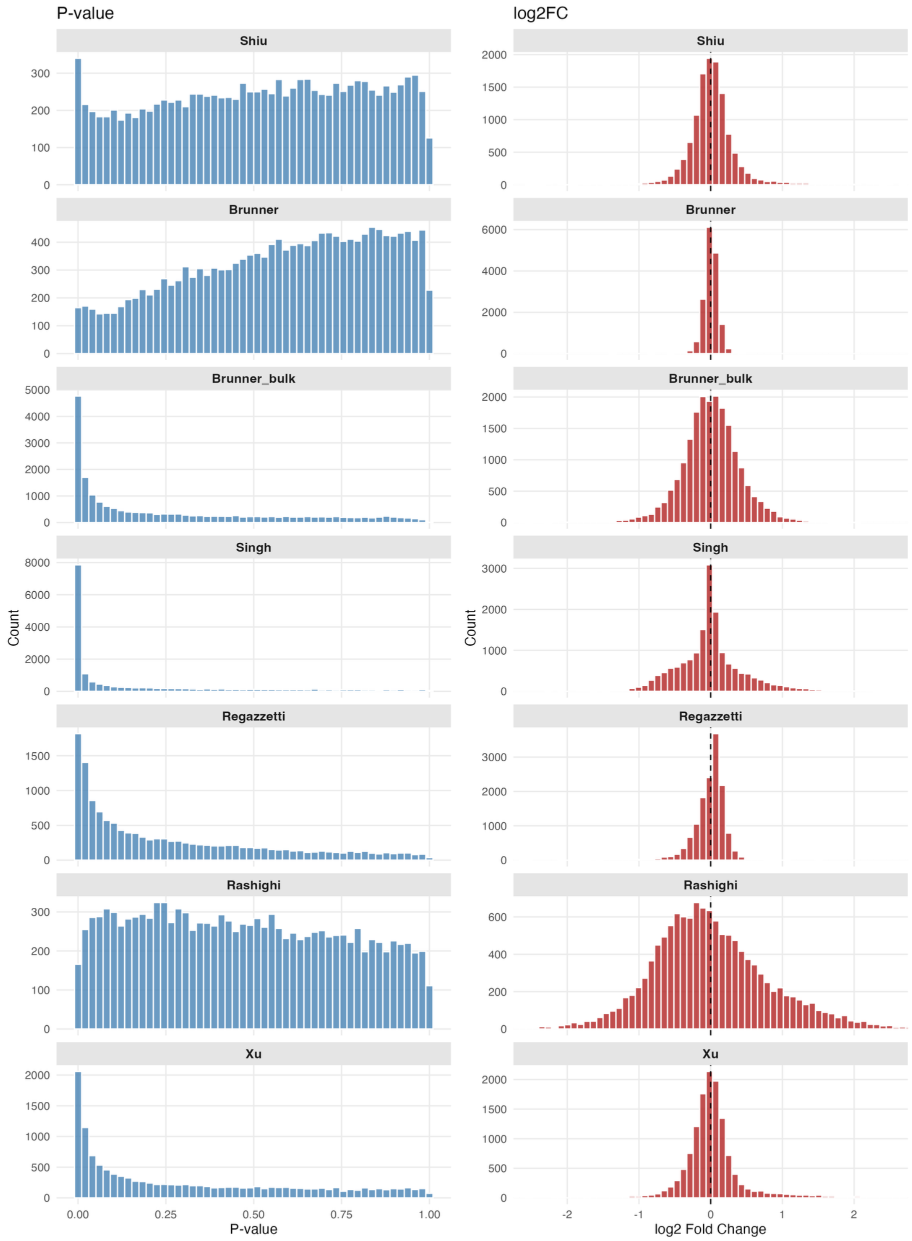


*Figure S1*

P-value distributions (left) and $log_2$ fold change distributions (right) for each study. Studies with larger sample sizes (Singh, Brunner bulk) show enrichment for small p-values, indicating stronger statistical power. Shiu and Rashighi show nearly uniform p-value distributions reflecting limited power. $Log_2$ fold change distributions are centered near zero with study-specific variance; Brunner pseudobulk has the narrowest distribution, reflecting conservative effect size estimates from the small sample sizes.

#### Figure S2. Gene Overlap Across Studies (Upregulated)

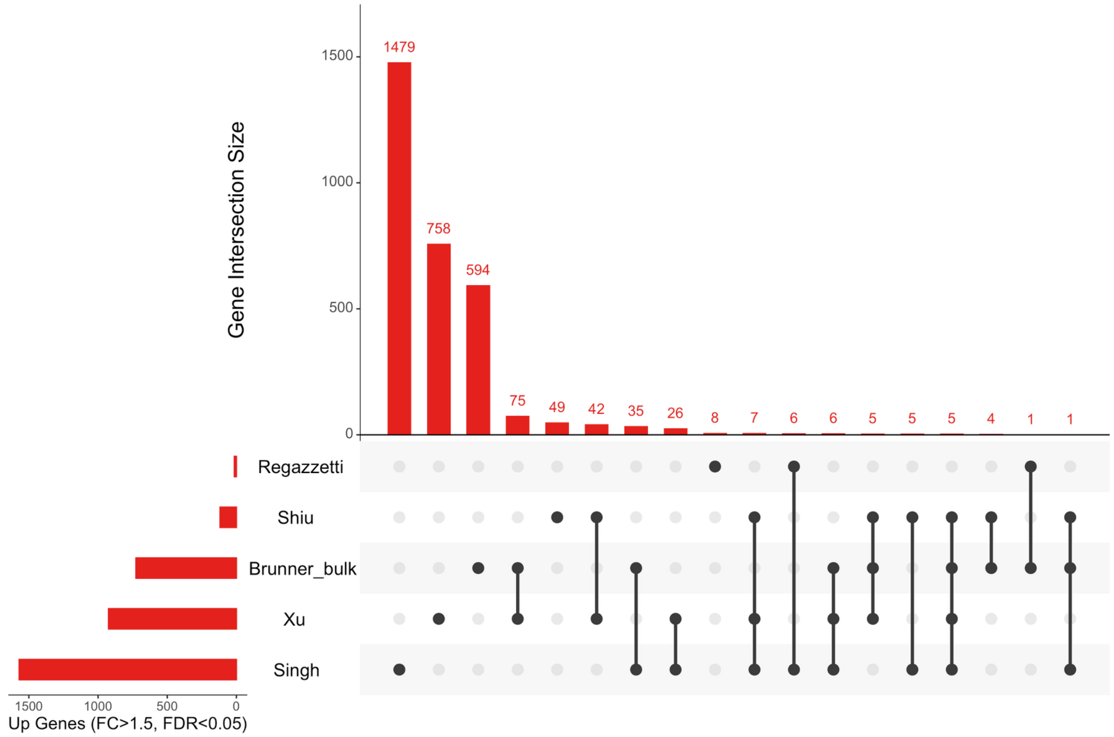


*Figure S2*

UpSet plot showing intersection sizes for upregulated genes (FDR < 0.05, FC > 1.5) across studies. Limited overlap is observed, with most genes detected in only one or two studies.

#### Figure S3. Gene Overlap Across Studies (Downregulated)

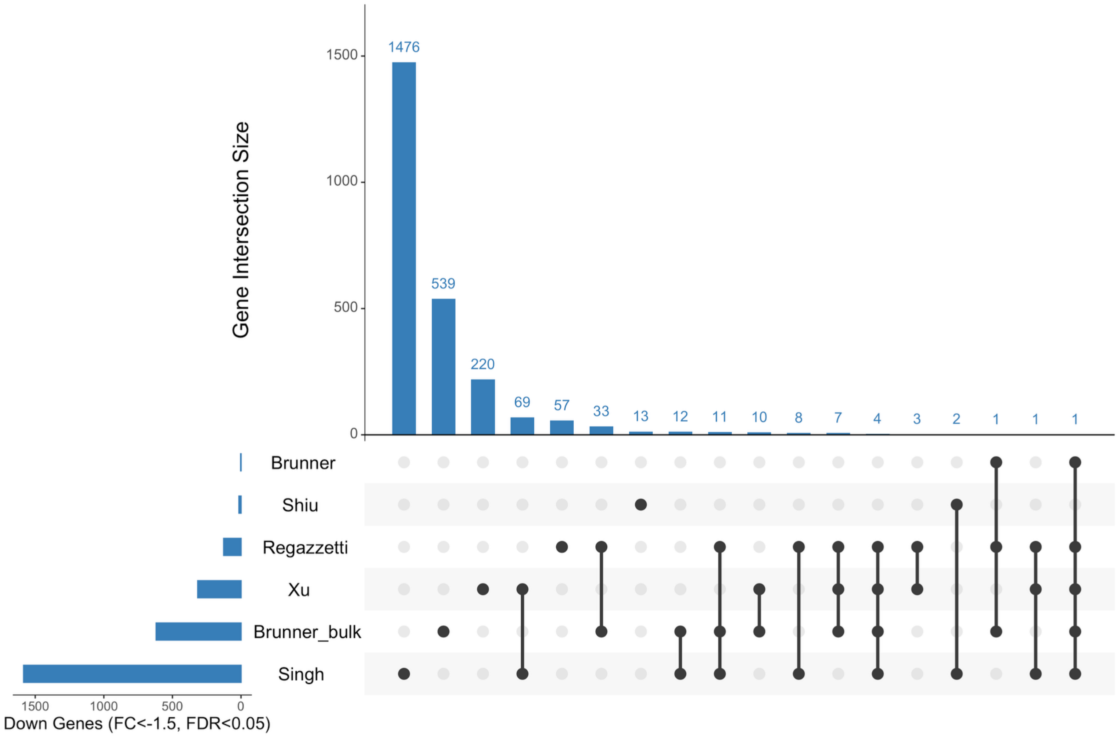


*Figure S3*

UpSet plot showing intersection sizes for downregulated genes (FDR < 0.05, FC > 1.5) across studies. Downregulated genes show better overlap than upregulated genes, with melanocyte markers appearing across multiple studies.

#### Figure S4. Cross-Study Effect Size Correlation

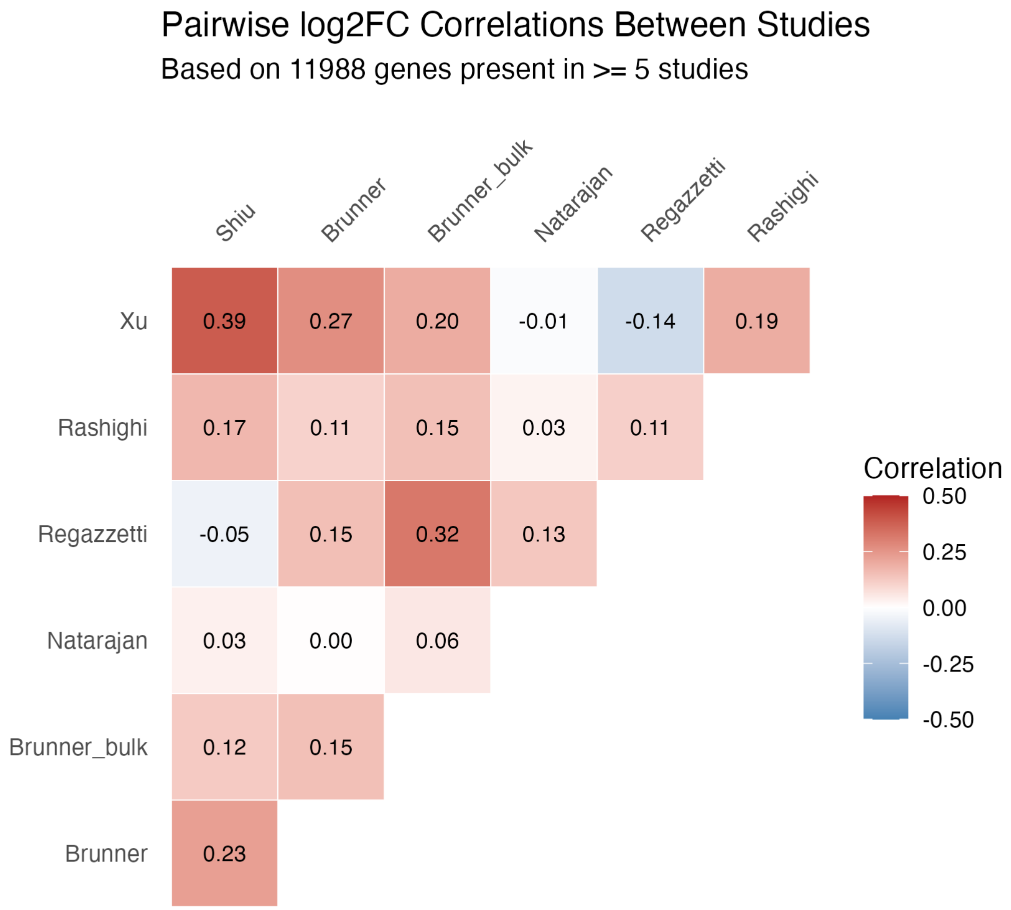


*Figure S4*

Pearson's correlation coefficient of $\log_2$ fold changes for 11,988 genes present in at least 5 studies. Correlations are generally low (range: −0.14 to 0.39), indicating substantial heterogeneity across studies and platforms. The highest correlation is between Shiu and Xu (r = 0.39), both pseudobulk datasets. Low cross-study correlation justifies rank-based meta-analysis over traditional effect-size pooling methods.

#### Figure S5. Rank Agreement Across Studies (Downregulated)

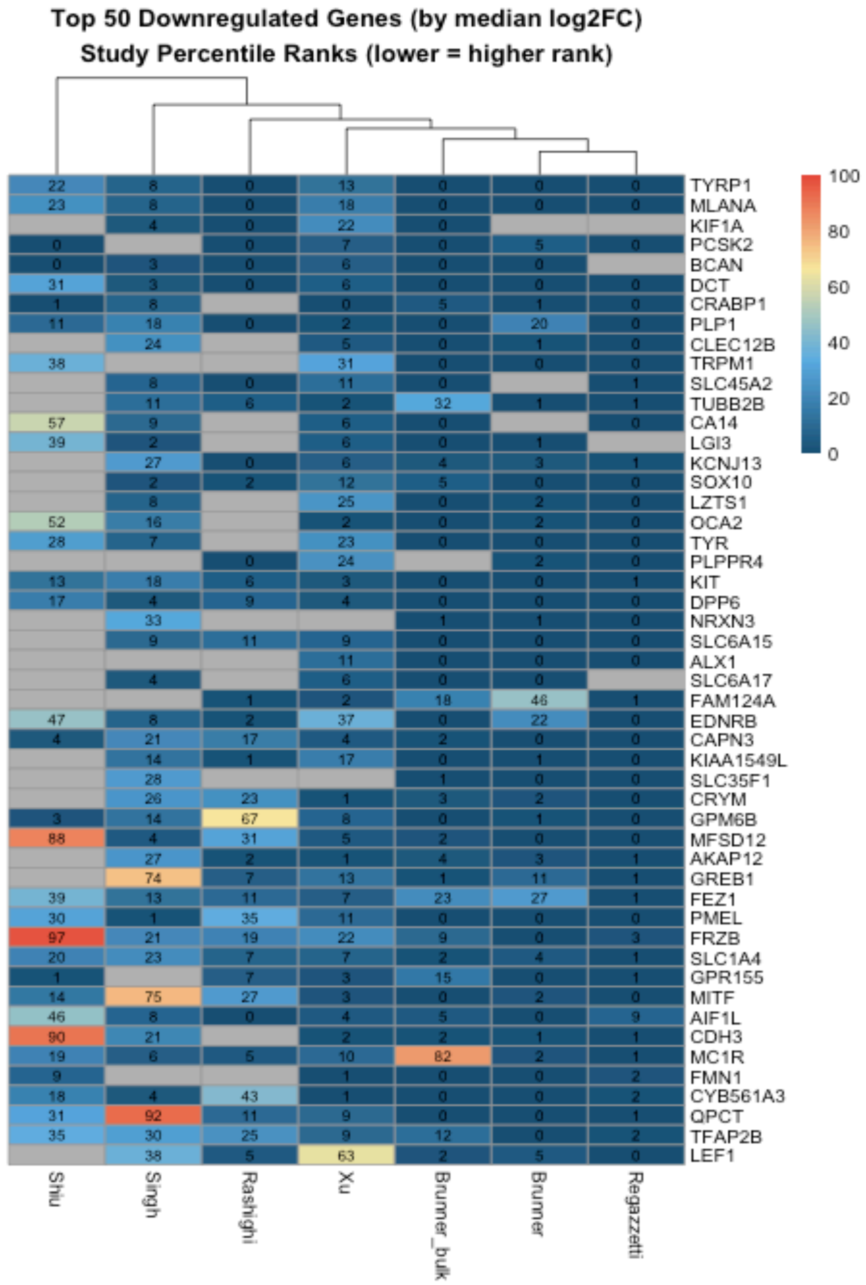


*Figure S5*

Heatmap showing study-specific percentile ranks for the top 50 downregulated genes ordered by median $log_2FC$. Lower percentile (orange/red) indicates a higher rank within that study. Despite divergent DEG counts across studies (0 to 3,154), top-ranked downregulated genes show consistent ranking across platforms. Melanocyte markers (TYRP1, MLANA, KIT, DCT) rank in the top percentiles across nearly all studies. The results from the Shiu dataset differ from those of the other studies.

#### Figure S6. Rank Agreement Across Studies (Upregulated)

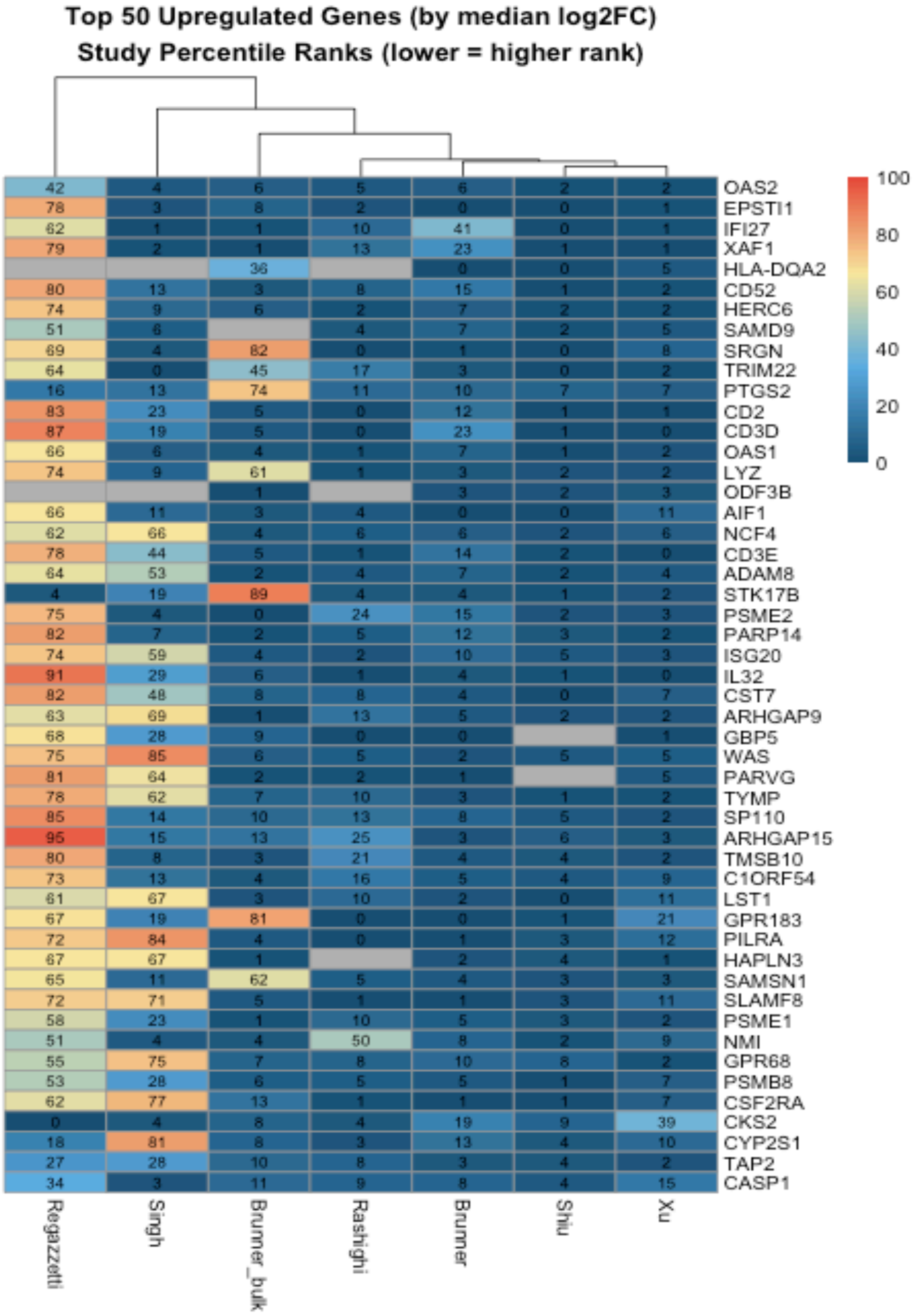


*Figure S6*

Heatmap showing study-specific percentile ranks for the top 50 upregulated genes ordered by median $log_2FC$. Lower percentile (orange/red) indicates a higher rank within that study. Upregulated genes exhibit more heterogeneous ranking patterns than downregulated genes, reflecting variable immune cell capture across platforms. The Regazzetti dataset shows weaker immune-response gene activation.

#### Figure S7. Leave-One-Out Robustness Analysis

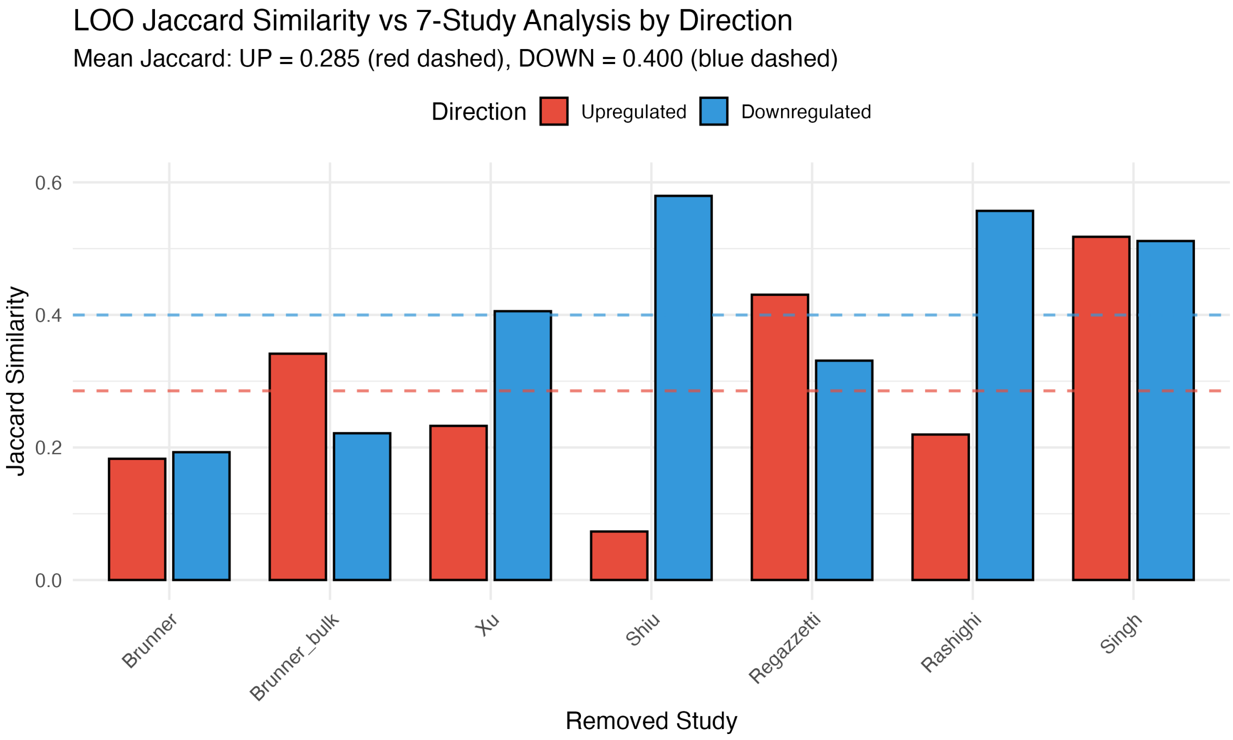


*Figure S7*

Jaccard similarity between leave-one-out (LOO) DEGs and those from the full 7-study analysis, separated by direction. Downregulated genes show consistently higher stability (mean Jaccard = 0.40) than upregulated genes. Removing the Shiu study causes the most dramatic loss of upregulated genes (Jaccard = 0.073), while downregulated genes remain largely intact (Jaccard = 0.58).

#### Figure S8. Over-Representation Analysis

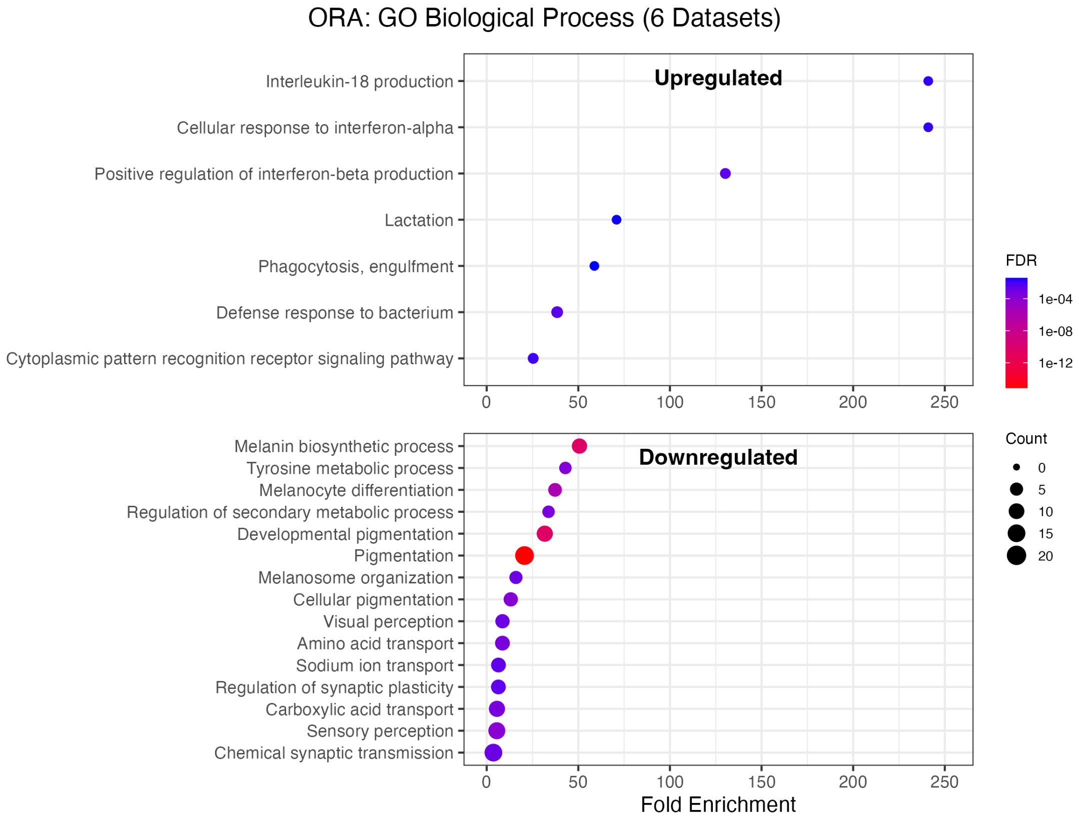


*Figure S8*

Over-representation analysis (ORA) dotplot showing fold enrichment for GO Biological Process terms among upregulated (top) and downregulated (bottom) genes (FDR < 0.05 from RRA). The 6 upregulated genes are enriched for interferon and antiviral response terms; the 108 downregulated genes are strongly enriched for pigmentation, melanin biosynthesis, and melanocyte development.

#### Figure S9. Shiu Study Investigation

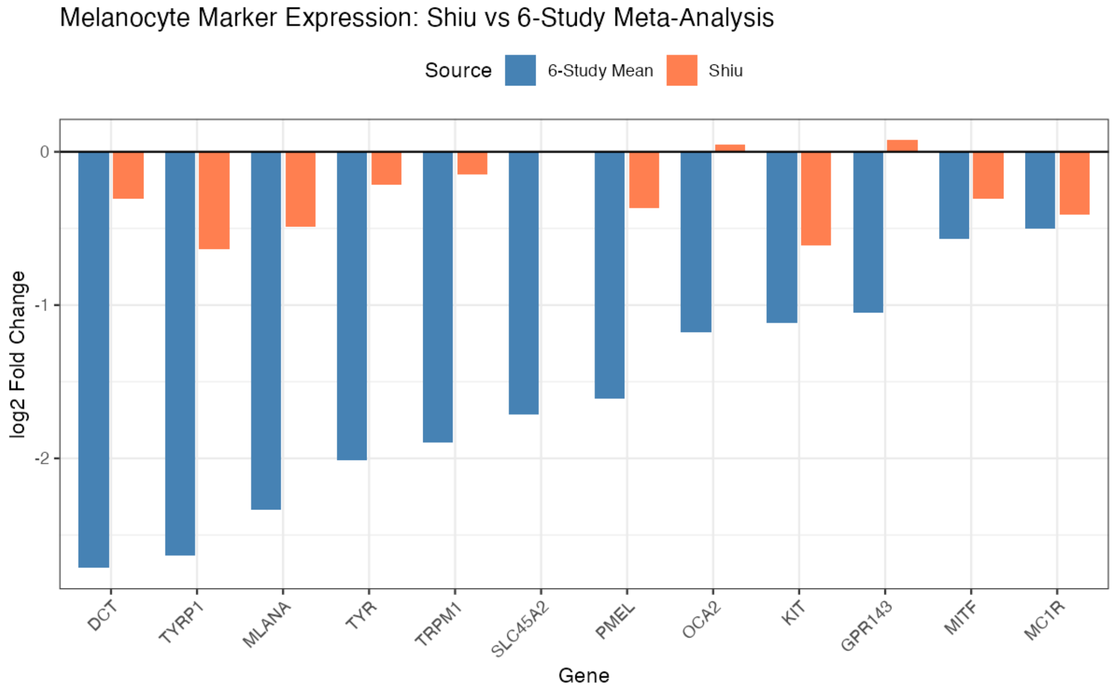


*Figure S9*

Comparison of melanocyte marker $log_2FC$ between Shiu and 6-study mean. While direction is concordant for most markers, effect sizes in Shiu are 3–5× smaller than the consensus, consistent with dilution of melanocyte signal by abundant immune cell transcripts in pseudobulk aggregation.

#### Figure S10. Average Expression of Downregulated Genes Across Cell Types

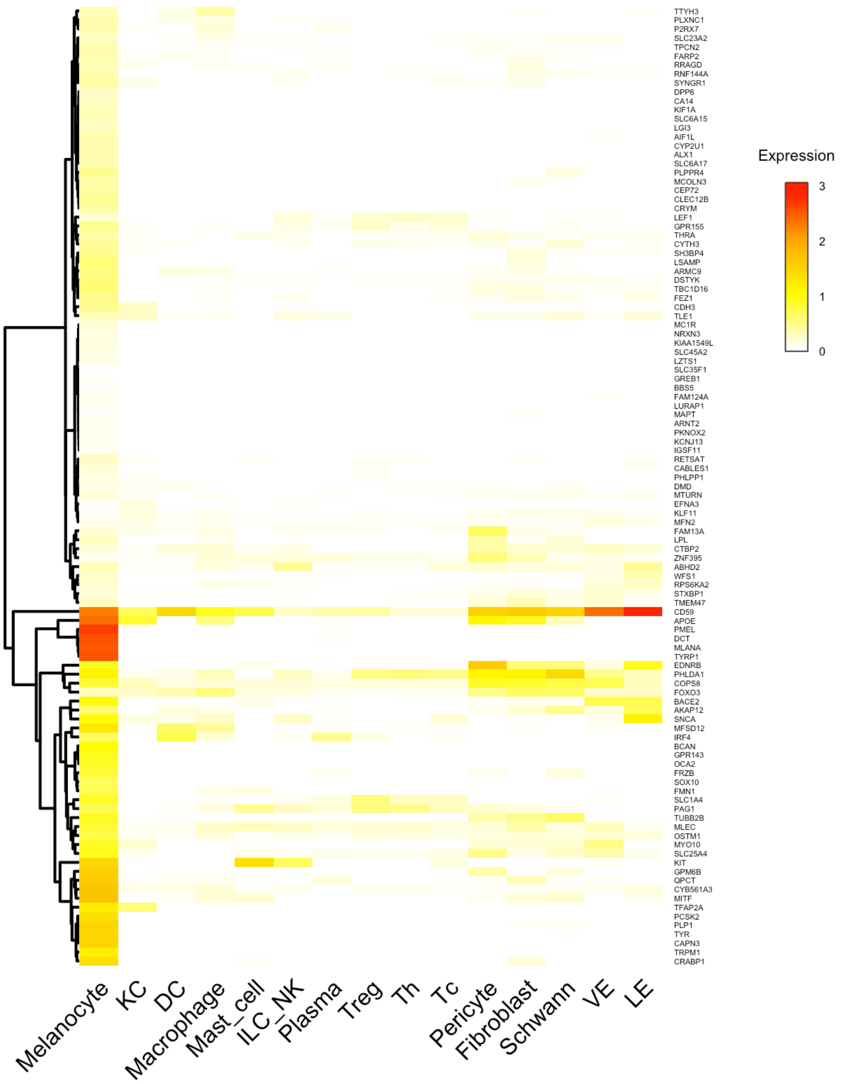


*Figure S10*

Mean log-normalized expression of meta-analysis downregulated genes (RRA FDR < 0.05) across cell types in the Reynolds healthy skin atlas. Hierarchical clustering reveals a melanocyte-specific cluster (bottom) containing canonical pigmentation genes (TYR, TYRP1, DCT, PMEL, MLANA) with expression levels 2–3 log units higher in melanocytes than other cell types. This complements the percent detection analysis (Figure 5B) by demonstrating that melanocyte-specific genes are not only more frequently detected but also expressed at substantially higher levels in melanocytes.
